# Harnessing routinely collected data for the evaluation of early years interventions: insights from a scoping review of evaluation studies

**DOI:** 10.1101/2025.10.29.25338970

**Authors:** Kate E. Mooney, Hollie Henderson, Kelly Hollingsworth, Sarah L Blower, Jenna Graham, Rina Davidson, Farwa Batool, Dea Nielsen

## Abstract

**Introduction:** Routinely collected data (RCD) describes data about individuals that is documented routinely in practice, typically in electronic medical, educational, and service records and registries. Utilisation of RCD for research can overcome limitations with traditional study designs, such as participant burden, attrition and disappointment bias. A systematic understanding of if (and how) RCD is being used to evaluate early years interventions is lacking.

**Objectives:** To scope the literature on how RCD is being used to evaluate the effectiveness of early years interventions being delivered in the UK.

**Methods:** The study protocol was registered online (osf.io/cug36). Included studies were interventions delivered to expectant parents and parents of children aged 0-5 years. Studies had to use a quantitative measure from RCD as an outcome. The study scope was limited to the UK, from January 2000 to November 2024. Completed studies and study protocols were eligible, encompassing both grey literature and peer reviewed sources. Medline, Psycinfo and Embase databases were searched via Ovid.

**Results:** 31 unique studies published 2009-2024 had used or were planning to use RCD as an outcome in an evaluation of an early years intervention. Most studies measured more than one outcome type, and the most common were birth outcomes and child education. Many studies noted limitations of using RCD, particularly being limited by the measures available in RCD, and fewer noted strengths.

**Conclusion:** Whilst the use of RCD expands evaluation opportunities, researchers are limited by what measures are available in RCD. We make four specific recommendations to improve the use of RCD in early years evaluations. With careful consideration, RCD can provide powerful insights regarding the impacts of early years interventions. This scoping review reveals an emerging methodology that may have increasing importance for evaluating early years policies and interventions but remains constrained by data availability and quality.

## Introduction

Providing all children with the best start in life is a high priority recommendation highlighted by UK government policies, showing a clear political commitment to early years support and investment (1). Early intervention delivered to children and their caregivers during pregnancy and in the first five years of their child’s life could prevent the onset of poor parental and child outcomes, and mitigate the potential personal, familial, and societal costs of longer-term negative outcomes (2). Early interventions can impact a range of caregiver and child outcomes, from physical health (e.g. birth outcomes, breastfeeding), wellbeing (i.e. maternal mental health, child socioemotional development), as well as developmental, cognitive and linguistic skills at the level of the child (i.e. early educational measures). Despite the potential benefits of intervening early being well documented (3), there remains a lack of robust evidence for the short and long-term effectiveness of interventions for parents of children in the critical early years of life (2,4).

This lack of robust evidence for such interventions may be due to the numerous challenges with evaluating them. Where individual services to support parents exist within local authorities, service providers may be reluctant to engage in randomised studies, as they prioritise providing support to those who need it most. Service providers may understandably be reluctant to use a randomised design because it could mean denying a potentially beneficial intervention to a family in need (5–7). Randomised studies that evaluate such interventions may also suffer from limited representativeness and generalisability, as parents who are at the point of ‘readiness to change’ are unlikely to agree to take part in a study in which they might be randomised not to get help, resulting in recruitment bias (8). Furthermore, such studies are often susceptible to recruitment challenges, particularly relating to a high number of participants being lost to follow up (9), especially when exploring long term effects of the intervention. For example, even with multiple incentives to support low-income mothers to remain in a parenting intervention, 41% of mothers were lost to follow up (10).

A potential solution to some of these challenges is to use routinely collected data (RCD), which describes data that are collected about individuals when they interact with public services, such as health, education and social care services, as part of their service delivery and practice (11,12). RCD can be used in studies examining the impact of factors such as early interventions, on parent and child outcomes (13,14). Hence, a solution to the issues with evaluating early years interventions is to instead utilise the data which are already, in theory, universally and routinely collected in parent and child records, overcoming limitations with recruitment and generalisability. RCD at a population level can also enhance generalisability and allow better comparison to similar populations (7). RCD enables both short and long-term outcomes to be obtained, which reduces the participant burden of extra data collection, and potentially reduces attrition and disappointment bias. This can be applied for both randomised and non-randomised designs, which means follow-up periods can be extended for randomised studies, and allows the realisation of observational studies of interventions. The use of non-randomised designs also reduces the ethical concern for the service regarding randomisation; as no participant who desires the intervention will later be randomised to a control group. However, it is also important to be aware of the limitations of non-randomised studies for making causal claims about the impacts of interventions (15).

It is important to note that since RCD is not designed for research purposes, its use for evaluating early years interventions must be considered with limitations in mind. One limitation is that the collection of RCD varies within the nations of the UK. For instance, whilst the Ages and Stages Questionnaire (ASQ) is a routinely used measure of child development as part of the Healthy Child Programme (16), the Schedule of Growing Skills (SOGS) is used in Wales (17). These differences limit the scope for evaluations across multiple nations. Previously noted concerns about RCD include the relevance of the RCD to the specific intervention, and the completeness and quality of the data itself (18–20). Regarding the relevance of the RCD, the effects of specific programmes may risk going unobserved if the routinely collected outcome lacks sensitivity to specific changes (19,20). A systematic review of the challenges and strategies for using RCD in research highlighted residual confounding, misdiagnosis, misclassification, and missing data as concerns (21).

New initiatives are facilitating access to routine data at both local (22,23) and national levels (12,24,25) for research purposes. For instance, RCD has been used to create Scotland’s first administrative child cohort, linking records for over 198,483 mother-child pairs (24), and England’s Education and Child Health Insights from Linked Data Mother-Baby (ECHILD-MB) cohort was created by linking 13.6 million baby records to mothers (25). These datasets can potentially be used to evaluate early years interventions, where information about intervention exposure has been routinely collected, or where population wide policies or interventions can be evaluated via natural experiments (15).

### Present study and rationale

Evidence for successful early years interventions is lacking, despite a renewed policy interest in providing all children with the best start in life (1,3). Despite its limitations, utilising RCD as an outcome could overcome several evaluation challenges. Hence, a systematic understanding of what components of RCD are being, or have been, used to evaluate early years interventions and/or policies, is crucial for planning of future evaluations. This may enable identification of areas where improvements to existing RCD infrastructures may be required, and identification of RCD that is being underused. Given the geographical variability of what and how data are collected between countries, it is important to consider these questions within the specific area of interest, in this case the United Kingdom.

This scoping review therefore aims to gain an overview of both planned and completed studies that have used RCD as an outcome in evaluations of early years interventions (delivered either during pregnancy, or up child age 5-years-old). Our focus is on universal and targeted interventions with a social and behavioural component, rather than interventions or products to treat specific conditions. The scoping review question is:

1. ***How is routine data being used to evaluate the interventions delivered in the early years in the UK?***

## Methods

A preliminary search of PROSPERO, the Cochrane Database of Systematic Reviews and JBI Evidence Synthesis was conducted and no current or underway systematic reviews or scoping reviews on the topic were identified. This scoping review was conducted in accordance with the JBI methodology for scoping reviews (26), and reported using the PRISMA Extension for Scoping Reviews (PRISMA-ScR) (27) (Appendix 1). The protocol was registered at https://osf.io/cug36/.

### Eligibility criteria

The ‘Population, Concept, Context’ framework was used to describe the eligible studies. Broadly, we focused on studies that included interventions delivered to caregivers with a 0-5-year-old child, and/or interventions delivered directly to children aged 0-5 years. The concept of the interventions was broad and could be any kind of intervention and/or policy that was delivered when parents were pregnant, or when the child was aged 0-5-years-old. The context was UK only, as the purpose was to inform the use of routine data use in intervention evaluations within UK contexts. Studies published since 1st January 2000 were included, to ensure that any results are relevant to modern data systems.

Table 1 describes the framework and eligibility criteria. A more detailed table with exclusion criteria for guiding the screening process was published with the study protocol (see osf.io/cug36/). At abstract screening, we included any studies that: (a) were interventions and used a quantitative design, (b) were about parents/children (aged 0-5-years-old) and (b) were in the UK, or it was not clear where the study took place.

**Table 1.**
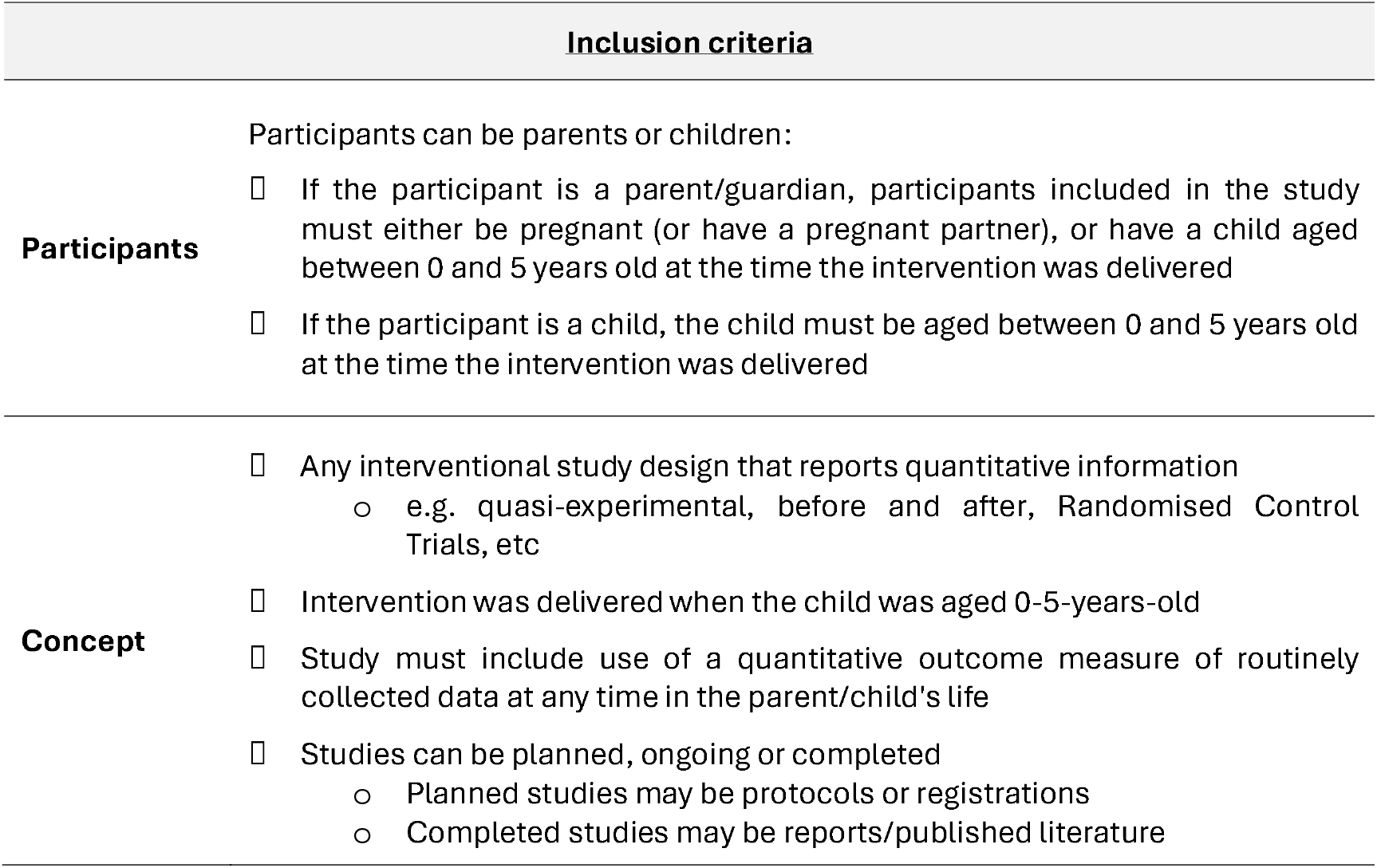
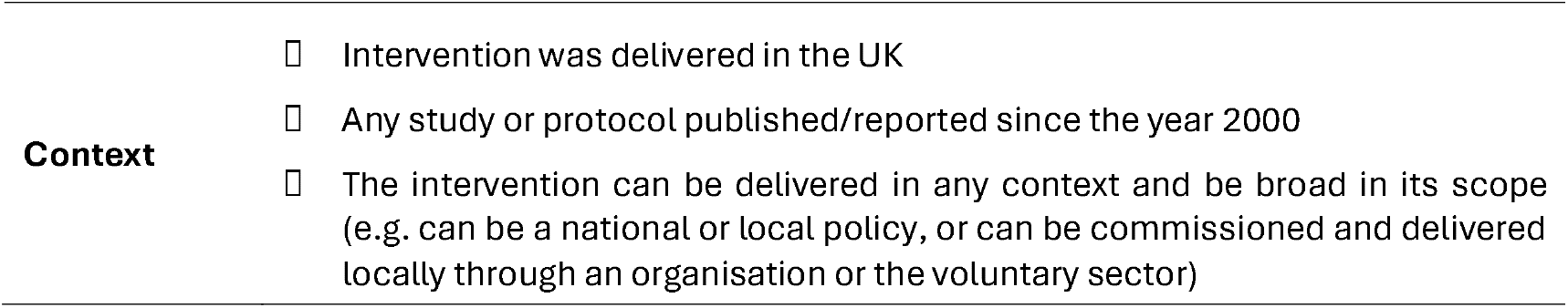
Participant, Concept, Context (PCC) Framework and eligibility criteria.

**Table 2.** Included studies characteristics.

| **<u>Characteristic</u>** | **<u>N (%)</u>** |
| --- | --- |
| **Study type (n=32 studies)** |  |
| Protocol | 12 (38) |
| Published study | 20 (62) |
| **Study design (n=31 unique studies)** |  |
| Observational studies (inc. natural/quasi-experimental) | 22 (70) |
| Randomised trial | 9 (29) |
| **Geographic location of study (n=31 unique studies)** |  |
| England |  |
| *National level* | 8 (26) |
| *Specific place in England* | 12 (39) |
| Scotland | 7 (23) |
| Wales | 2 (6) |
| Multiple nations | 2 (6) |
| **Intervention focus (n=31 unique studies)** |  |
| Antenatal programme | 9 (29) |
| Service/policy | 7 (23) |
| Smoking cessation | 3 (10) |
| Breastfeeding | 3 (10) |
| Parenting programme | 3 (10) |
| Voucher programme | 2 (6) |
| COVID-19 | 2 (6) |
| Weight | 2 (6) |
| **Outcome data (n=31 unique studies)** |  |
| Parent | 10 (32) |
| Child | 11 (35) |
| Both | 10 (32) |
| **Time period of latest outcome data (n=31 unique studies)** |  |
| Pregnancy | 2 (6) |
| Birth | 7 (23) |
| Post birth | 10 (32) |
| Up to child age: |  |
| *1 year* | 1 (3) |
| 3 years | 3 (10) |
| 4 years | 1 (3) |
| 5 years | 4 (13) |
| 6 years | 1 (3) |
| 7 years | 2 (6) |

### Types of Sources

Peer reviewed studies and grey literature (including reports, websites, and conference abstracts) were both eligible for inclusion. We included studies at any stage from study protocols, interim findings, to study reports of outcomes.

### Published literature search strategy

The full search is provided in Appendix 2. The words associated with the population, concept and context of the review were used to guide the search terms. The National Institute for Health and Care Excellence (NICE) filters were used to identify studies conducted within the United Kingdom (28). Medline, Psycinfo and Embase were searched via Ovid for peer reviewed literature published between January 2000 and 13th November 2024.

### Grey literature search strategy

Grey literature searches took place in November 2024. We targeted specific websites relevant to our topics, namely National Institute for Health Research (NIHR), Education Endowment Foundation (EEF), Early Intervention Foundation, National Foundation for Education Research (NFER), ISRCTN (https://www.isrctn.com/),Clinicaltrials.gov, GOV.UK (https://www.gov.uk/search/all),and GOV.wales (https://www.gov.wales/). Records of websites searched, number of potentially relevant items, and number scanned were kept in Microsoft Excel (29).

### Evidence selection

Titles and abstracts were uploaded to Covidence for screening. A piloting process took place with 100 studies, and agreement between all reviewers was reviewed. After piloting, amendments were made to the inclusion criteria to improve the clarity of the included studies. All titles and abstracts were screened by two reviewers according to the criteria described above for abstract screening. Cohen’s Kappa scores were reviewed regularly as a team, since lower than desired agreement (<.60) occurred between some pairs of reviewers. Regular meetings and reviews took place with the whole team to resolve disagreements, and Cohen’s Kappas improved. After abstract screening, the full texts were screened and agreed by two reviewers (DN or KEM).

### Data extraction

Data was extracted from eligible papers by three reviewers (RD, JG, KH). A proportion of 20% of selected articles was second checked by KEM to ensure accuracy. Data included specific details about the participants, concept, context, study methods and key detail regarding the outcome measurement and source of RCD. Discussions of studies were examined to extract a list of specific strengths and limitations associated with using RCD.

### Data analysis and presentation

A full list of studies and characteristics is provided in Appendix 3. The included studies were summarised in terms of their study types, designs, intervention types, and outcomes used including the type of outcome, timing, and whether the outcome was reported at the parent or child level. Data were charted by these study characteristics, and bar graphs created where they provided additional insights.

## Results

Figure 1 presents the flow of included studies. There were two studies using the same data source, but they contained different periods of follow up routine data, hence, we retained these two studies in the review. The studies are (1) a protocol of a randomised study to evaluate the Family Nurse Partnership (30) and the results from the routine data follow up from that study (31).

**Figure 1.**
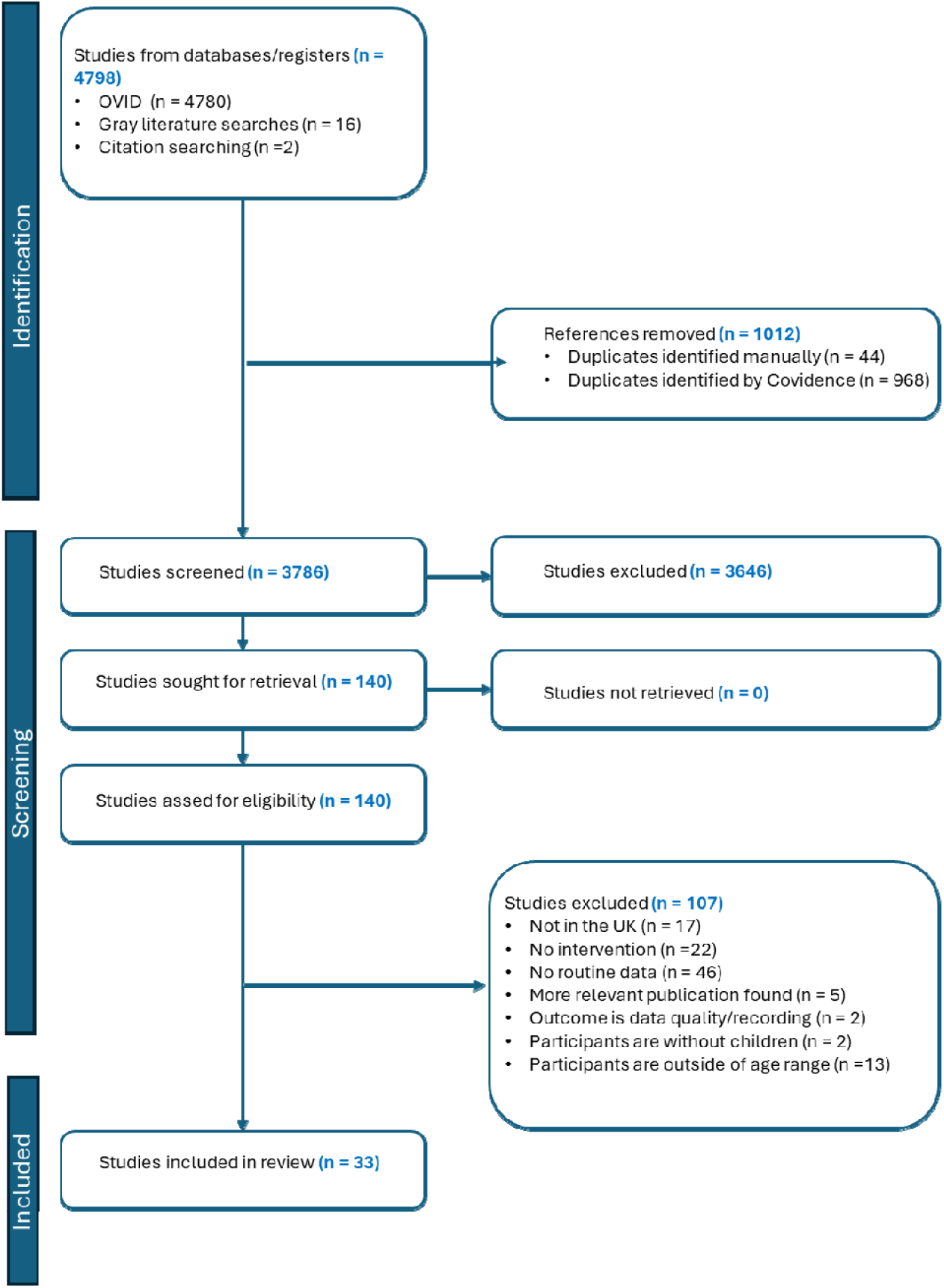
PRISMA flow diagram for scoping review process Search results

**Figure 2.**
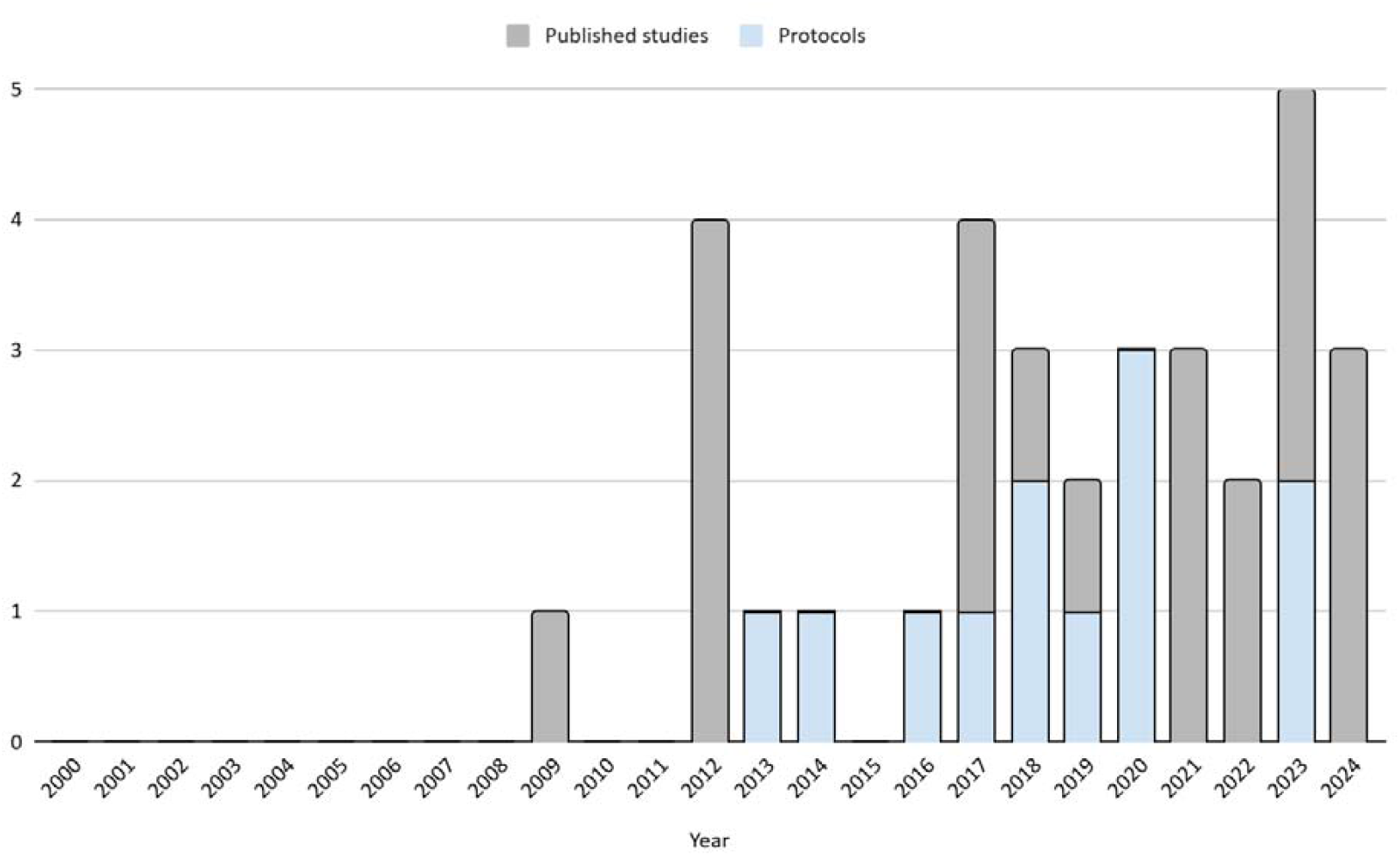
Number of studies published per year

### Study characteristics

Table 3 summarises key characteristics of the included studies, and the extracted data for all studies is provided in Appendix 3. This information is used in the narrative synthesis below.

### Narrative synthesis

#### Description of studies

Out of 32 studies, 20 were published studies with results (62%), and 12 were study protocols (38%). Out of 31 unique studies, most (n=22, 71%) were observational (ie. non-randomised interventional studies). The remainder were randomised control trials (n=9, 29%) (30,32–40).

The first eligible study was published in 2009. Generally, the number of published studies eligible for our review has increased with time, with the most being five studies in 2023, which included two protocols and three published studies.

#### Intervention type

Most (n=9, 29%) were perinatal interventions, meaning that they were broad perinatal support interventions that started antenatally, with some continuing postnatally. This included programmes such as the Family Nurse Partnership (30,38,41,42), “Pregnancy Circles” (35,36), and “Baby Steps” (43). The second most common category were service and/or policy evaluations (n=7, 23%). This included a broad remit of intervention types ranging from, for example, the availability of Community Perinatal Mental Health teams in local areas (44), to an evaluation of the impact of a Growth Assessment Protocol implemented in maternity (40). The other categories were smoking cessation in pregnancy (n=3, 10%), breastfeeding support programmes (n=3, 10%), parenting programmes (n=3, 10%), voucher schemes (n=2, 6%), weight programmes (n=2, 6%), and the effect of COVID-19 policies (n=2, 6%).

#### Routinely collected outcomes

*Note: studies may have measured >1 outcome, so double counting of studies occurs*.

There was an even spread of studies which focused on only parent outcomes (n=10, 32%), only child outcomes (n=11, 35%), and both parent and child outcomes (n=10, 32%). Figure 3 shows that we categorised outcomes into 14 broad categories. More than half of the studies measured more than one outcome type (n=19, 61%), with the most being six different outcome types in one study (41). The most common category was birth outcomes (n=12, 39%), which included a broad range such as gestational age, birth type, and stillbirths. These studies mentioned use of maternity electronic patient records (45), Hospital Episode Statistics (HES) (46), and routinely collected NHS data (47). The next most common was child education and/or development outcomes (n=7, 23%), with all but one of these studies mentioning use of the Early Years Foundation Stage Profile (EYFSP) linked via the National Pupil Database (31,39,41,42,48,49). The one study which did not use the EYFSP was a protocol for evaluating the impact of the universal health visiting pathway in Scotland on child developmental concerns at age 27-30 months, though does not state the exact measure used (50).

**Figure 3.**
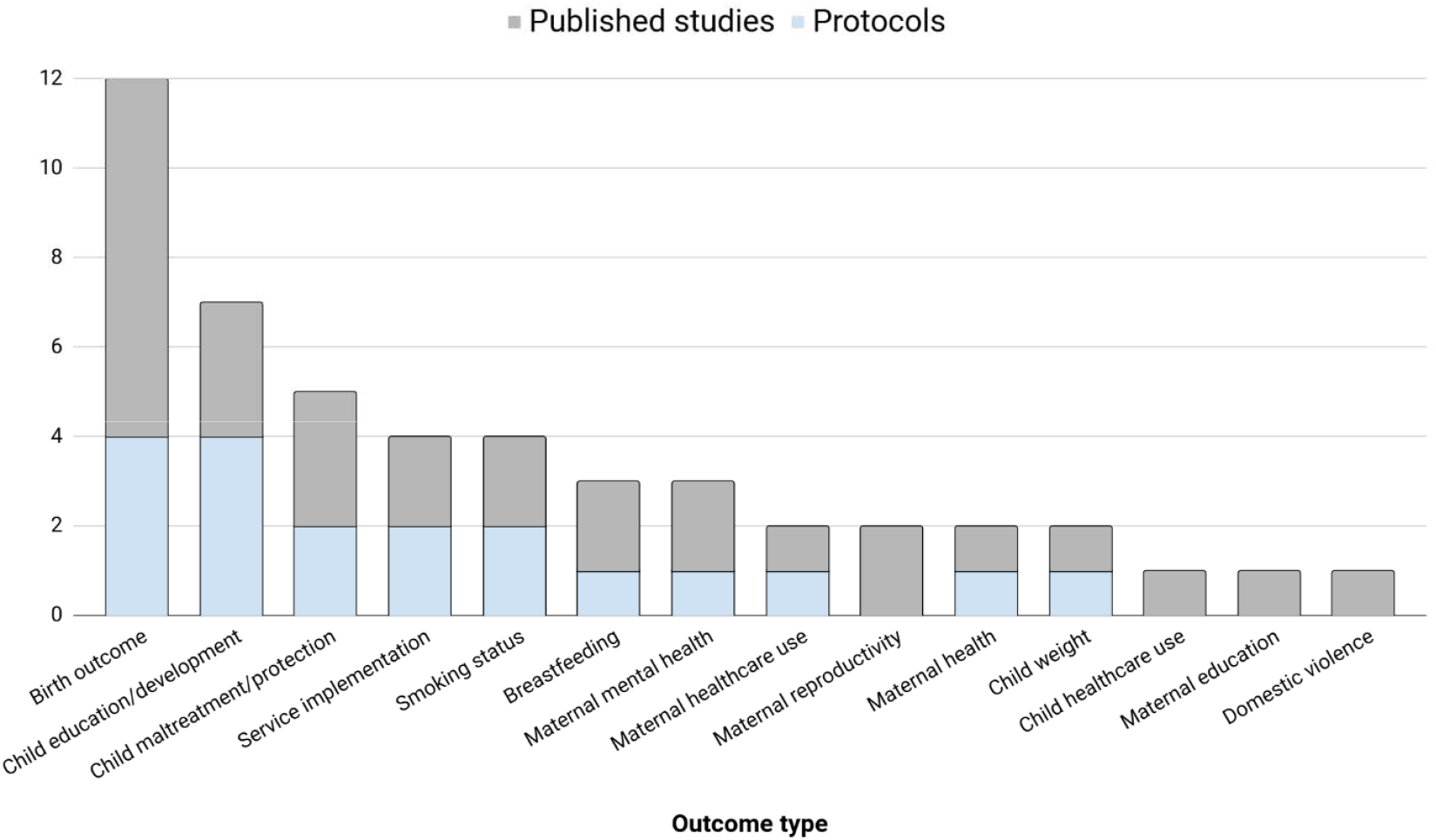
Outcome types measured in all studies, separated by protocol and publishe studies

The next most common was child maltreatment (n=5, 16%), with one study linking this via child protection status in administrative data (51), two using social care data (41,50), one using local authority child protection register (52), and one ascertaining this via Child in Need (CIN) status via the National Pupil Database (42). The next most common outcome was service implementation (n=4, 12%), which related to the proportion of women attending antenatal bookings (35,36), child health reviews conducted by GPs (53), and service use of a community perinatal mental health team (44). The next most common was smoking status (n=4, 14%), recorded in maternity records during pregnancy (50,54–56).

We categorised studies into the timing of their most recently routinely collected outcome. The most common period was up to 3 months post birth (n=10, 32%). This included length of stay in hospital (40,46) and readmission (46), breastfeeding outcomes (32–34), survival for the parent and/or baby (40,57). The next most common follow up period was at birth (n=7, 23%). Very few studies examined outcomes beyond 5 years (n=3, 10%).

#### Limitations and strengths noted in studies

Appendix 3 presents the list of strengths and limitations per study, and we summarise the findings here. Most of the studies (n=20, 62% of 32 studies total) mentioned one or more specific limitations of using RCD. A common limitation noted by nine studies was that the choice of outcome measurement was limited by what was available in routine data (31,43,48–51,53,56,58). Another limitation was the high level of missing data (44,52,54,59). Other common limitations noted were the length of time to gain permissions to access and receive data (41,48), and concerns in the accuracy of the data (32,34,44,60).

Fewer studies mentioned strengths of using RCD (n=9, 28% of 33 studies total). Several studies noted the increased sample size achieved through using RCD, noting whole population coverage(32,35,45,57), low missing data, low loss to follow up (33), and improved power and/or greater precision in estimates (44,54). Two studies noted that using RCD is cost-effective, highlighting the low burden for participants (42,47). Two studies noted the relevance of the outcomes to clinical practice and/or policy making decisions (45,49). One study noted the minimisation of bias in self-reporting, and the potential to follow up participants over time (42).

## Discussion

### Summary of studies

We identified a total of 31 unique studies published between 2000-2024 that had used or were planning to use RCD as an outcome to evaluate an early years intervention. The majority were publications with results (64%), with the remainder being study protocols (38%). The studies were predominantly based in England (65%), followed by Scotland (23%), Wales (6%), or multiple nations (6%). The few studies using data from >1 nation may suggest a challenge in harmonising RCD across the UK’s devolved nations, since these nations operate separate early years services and hence separate data collection systems (16,17). The first eligible study was published in 2009. The number of eligible studies has increased over time, with the highest number being five in 2023. This could reflect both a developing field due to an increased policy demand for effective early years interventions (1,61), and an increase in data linkage research and capabilities (13,25).

Most studies employed observational designs (n=22, 71%). Observational studies were likely most frequent due to the nature of RCD, which is that it is primarily collected for purposes other than research (62,63). RCTs typically represent funded studies which do not usually need to rely upon RCD. The most common type of intervention were perinatal programmes (29%). This may reflect the availability of RCD in maternity records, meaning these programmes are readily evaluable using RCD on birth related outcomes.

### Outcomes in routinely collected data

The common use of both parent and child outcomes demonstrates the benefits of using RCD, as it can assess the dual impact often targeted by complex interventions delivered in the early years. Evaluations that measure both categories are best positioned to capture the full scope of an intervention’s potential effectiveness.

Most studies measured more than one outcome type via RCD. This highlights a strength of using RCD, as it has the capacity for comprehensive and multi-purpose evaluation with no additional data collection burden for participants or researchers. This aggregation may allow evaluations to move beyond a single “primary” outcome, providing a more comprehensive understanding of an intervention’s effects across various domains. As noted by many of these studies, use of RCD is highly cost-effective and low burden compared to collecting multiple different outcomes through bespoke surveys.

The most common outcomes were birth outcomes and child education/development. The commonality of birth outcomes may, again, reflect the availability of RCD in maternity records to be used in such evaluations. These birth outcome data indicators may be recorded with higher completeness than other RCD, because they are embedded into midwifery practice, and essential for clinical decision-making. These indicators therefore may serve as reliable, early indicators of intervention success, particularly for antenatal programs. The frequent use of the EYFSP to measure child development is likely due to it being a mandated assessment in England, meaning it has high coverage for this population at school entry (64). Use of the EYFSP may allow researchers to assess the long-term developmental impact of perinatal interventions, a key advantage over short-term studies. This would also reflect a key policy interest, with the UK Government target for 75% of children to have a Good Level of Development on the EYFSP by 2028 (1).

The routinely collected ASQ by health visitors at the 2-2 ½ year review completed in England’s Healthy Child Programme (16,65) was not indicated to be used by any studies, except potentially by one (50). The ASQ is a broad measure of a child’s early development and could be a targeted outcome for many interventions, and this suggests that this potential measure may be being underutilised. However, whilst the ASQ should be universally completed as a mandated assessment in England (16,65), recent studies have indicated variation in the completeness of the ASQ (66), which may explain its underuse as an outcome in evaluations.

Whilst many studies targeted the same outcomes, they differed in their descriptions of their data source. For instance, several studies utilised routine maternity data, however, it was challenging to determine if they had used the same data sources e.g. “maternity electronic patient records,” and “routinely collected NHS data”. These terms may describe the same underlying data source, or distinct datasets, and the variation in language makes this difficult to ascertain. The lack of standardisation in describing routine data sources presents a barrier for researchers seeking to use maternity data to evaluate early years interventions.

The timing of the outcome varied, with the most common being up to 3 months post birth (32%), at birth (23%), and at child age 5 years (13%). This indicates that most studies used RCD in the very earliest years of children’s lives, and fewer studies successfully leveraged RCD for medium-to-long-term follow-up (with only 3 studies examining outcomes post 5 years-old). A main benefit of RCD is the possibility for long term follow-up with minimal participant burden. It is possible that as access to RCD improves, more studies linking long-term RCD to evaluations will emerge.

### Strengths and limitations noted in studies

Most studies noted specific limitations in using RCD in their studies, with the most frequent being the limited choice of outcome measurement based on what was available in the RCD. This limitation highlights that crucial, but more nuanced, proximal outcomes (e.g., parental confidence, quality of parent-child interaction) that might typically be collected may be missed in a study relying upon RCD (19,20). Further, the measures collected in RCD may not be the most valid and reliable measures of specific outcomes. This could lead to an incomplete picture of an intervention’s effect, particularly if the mechanism of change is not directly measurable in RCD. Political interest in using population-based data for evidence-based decisions is crucial for developing new databases and data collection systems (7), hence the current Government Best Start policy (1) highlights the timeliness of this review.

Authors also noted concerns regarding accuracy of the data, and this should be investigated by researchers, for instance via studies which examine the implementation of RCD in local practice (67). These limitations reflect that RCD are not designed for research purposes, and its use should always be considered with its limitations in mind (18,21).

Fewer studies noted strengths (30% of 33 studies), which most frequently included an increased sample size and whole population coverage, and low missing data or low loss to follow up. RCD may address previously noted challenges with evaluating early years interventions, including that such interventions are subject to a high attrition rate (8,10). Altogether, the limitations and strengths highlighted a critical trade-off when using RCD to evaluate interventions: whilst the use of RCD addresses some challenges with evaluating early years interventions, researchers are also limited by what was available in RCD.

### Recommendations

1. **Policymakers to integrate further outcomes into RCD**. RCD currently misses key outcomes for early years interventions. Policymakers should mandate the integration of standardised, brief self-report measures (e.g., validated scales for parental mental health or confidence) into routine collection points (e.g., universal health visitor checks). This integrated data could be designed for both clinical and evaluation use.
2. **Researchers to communicate the use of RCD to services**. Many studies noted difficulties with accessing high quality and well completed RCD. Researchers must communicate the impact of RCD findings back to service providers. Demonstrating how this data informs intervention evaluation could improve service delivery, and boost motivation for services to ensure data completeness, accuracy, and accessibility.
3. **Researchers, services, and policymakers to develop a standardised terminology for describing RCD sources and access**. Descriptions of RCD sources and measures differed between studies. Clear, consistent descriptions of datasets would enable researchers to better understand the characteristics of different data sources, including their coverage, completeness, and the specific variables captured. A co-produced, standardised terminology could enable researchers to better navigate the process of obtaining data, and support more efficient use of RCD.
4. **Researchers and services to improve data linkage and utility of underrepresented outcomes**. Some key outcomes were underutilised, potentially indicating poor data quality and/or accessibility. Future work should be dedicated to understanding reasons for their underuse, and developing robust linkage mechanisms for these currently underused datasets.
5. **Researchers to continue to utilise RCD and bespoke collected outcomes to understand the effects of interventions**. RCD is not sufficient to be used on its own to evaluate interventions and policies. Hence, evaluations with bespoke data collection are still required. Data from these studies can be combined with outcomes nested in RCD where possible, to build understanding of the effects of early life interventions.

### Limitations

We did not formally assess the quality or risk of bias of the included studies. Consequently, we could not evaluate the quality of the RCD sources used or the potential impact of data quality on intervention evaluations. Since the use of RCD for research purposes is a rapidly evolving field, these review findings may require updating within a few years. Finally, although we made efforts to include grey literature, sources such as trial registrations and conference abstracts often lacked the detailed methodological information needed to confirm eligibility. This means the current review likely underestimates the volume of relevant studies that exist and may become more widely available in the near future.

## Conclusions

Altogether, this review has highlighted a critical trade-off when using RCD to evaluate interventions: whilst RCD opens up evaluation opportunities, researchers are also limited by what measures are available in RCD. The recommendations call for (1) policymakers to integrate relevant measures into routine data (RCD), (2) researchers to actively communicate findings to services to improve data quality; (3) stakeholders to standardise RCD source terminology and (4) researchers and services to improve data linkage for underused RCD. With careful considerations, RCD can provide powerful insights into the effectiveness of early years interventions, and researchers should continue to use it in combination with other available data. This scoping review reveals an emerging methodology that may have increasing importance for evaluating early years policies and interventions, but remains constrained by data availability and quality.

## Supporting information

Appendix 1 - PRISMA ScR Checklist

Appendix 2 - Search terms

Appendix 3 - Study data

## Data Availability

All data produced in the present study are available upon reasonable request to the authors.

## Acknowledgements

The authors of this study are grateful for the contributions from the Better Start Bradford Innovation Hub who made meaningful recommendations to the development of the protocol.

## Data availability

This scoping review did not include any primary data collection. All data produced in the present study are available upon reasonable request to the authors.

## Ethics statement

This study did not involve any primary data collection and hence did not require ethical approval. All data is extracted from publicly available studies.

## References

1. GOV.UK. Giving every child the best start in life - GOV.UK [Internet]. 2025 [cited 2025 Oct 16]. Available from: https://www.gov.uk/government/publications/giving-every-child-the-best-start-in-life

2. Hurt L, Paranjothy S, Lucas PJ, Watson D, Mann M, Griffiths LJ, et al. Interventions that enhance health services for parents and infants to improve child development and social and emotional well-being in high-income countries: a systematic review. BMJ Open [Internet]. 2018 Feb 1 [cited 2024 Apr 19];8(2):e014899. Available from: https://bmjopen.bmj.com/content/8/2/e014899

3. Heckman JJ. SCHOOLS, SKILLS, AND SYNAPSES. Econ Inq [Internet]. 2008 Jul 1 [cited 2025 Oct 17];46(3):289–324. Available from: 10.1111/j.1465-7295.2008.00163.x

4. Pidano AE, Allen AR. The Incredible Years Series: A Review of the Independent Research Base. J Child Fam Stud [Internet]. 2015 Jul 8 [cited 2022 Nov 21];24(7):1898–916. Available from: https://link.springer.com/article/10.1007/s10826-014-9991-7

5. Zimmerman FJ. Population Health Science: Fulfilling the Mission of Public Health. Milbank Quarterly. 2021;99(1):9–23.

6. Mooney KE, Welch C, Crossley K, Bywater T, Wright J, Dickerson J, et al. Is it feasible to nest a Trial within a Cohort Study (TwiCS) to evaluate an early years parenting programme? A Born in Bradford’s Better Start study protocol. Pilot Feasibility Stud [Internet]. 2024 Jan 30;10(1):19. Available from: https://pilotfeasibilitystudies.biomedcentral.com/articles/10.1186/s40814-023-01441-9

7. Morrato EH, Elias M, Gericke CA. Using population-based routine data for evidence-based health policy decisions: lessons from three examples of setting and evaluating national health policy in Australia, the UK and the USA. J Public Health (Bangkok) [Internet]. 2007 Dec 1 [cited 2025 Oct 21];29(4):463–71. Available from: 10.1093/pubmed/fdm065

8. Stewart-Brown S, Anthony R, Wilson L, Winstanley S, Stallard N, Snooks H, et al. Should randomised controlled trials be the “‘gold standard’” for research on preventive interventions for children? J Child Serv. 2011;6(4):228–36.

9. Hariton E, Locascio JJ. Randomised controlled trials—the gold standard for effectiveness research. BJOG [Internet]. 2018 Dec 1 [cited 2022 Nov 21];125(13):1716. Available from: /pmc/articles/PMC6235704/

10. Katz KS, El-Mohandes A, Johnson DMN, Jarrett M, Rose A, Gober M. Retention of low income mothers in a parenting intervention study. J Community Health [Internet]. 2001 [cited 2025 Sep 2];26(3):203–18. Available from: https://link.springer.com/article/10.1023/A:1010373113060

11. McDonald L, Lambrelli D, Wasiak R, Ramagopalan S V. Real-world data in the United Kingdom: Opportunities and challenges. BMC Med [Internet]. 2016 Jun 24 [cited 2025 Sep 2];14(1):1–3. Available from: https://link.springer.com/articles/10.1186/s12916-016-0647-x

12. Administrative Data Research UK. Data-driven change - ADR UK [Internet]. [cited 2025 Oct 17]. Available from: https://www.adruk.org/

13. Stewart E, Brophy S, Cookson R, Gilbert R, Given J, Hardelid P, et al. Using administrative data to evaluate national policy impacts on child and maternal health: a research framework from the Maternal and Child Health Network (MatCHNet). J Epidemiol Community Health [Internet]. 2023 Nov 1 [cited 2024 Jun 20];77(11):710–3. Available from: https://jech.bmj.com/content/77/11/710

14. Foust R, Hoonhout J, Eastman AL, Prindle J, Rebbe R, Nghiem H, et al. The Children’s Data Network: Harnessing the scientific potential of linked administrative data to inform children’s programs and policies. Int J Popul Data Sci [Internet]. 2021 [cited 2024 Jun 20];6(3). Available from: /pmc/articles/PMC9053132/

15. Skivington K, Matthews L, Simpson SA, Craig P, Baird J, Blazeby JM, et al. A new framework for developing and evaluating complex interventions: update of Medical Research Council guidance. BMJ [Internet]. 2021 Sep 30 [cited 2023 Feb 27];374. Available from: https://www.bmj.com/content/374/bmj.n2061

16. Shribman S, Billingham K. Healthy Child Programme. Pregnancy and the first five years of life. [Internet]. 2009 [cited 2024 Apr 10]. Available from: https://www.gov.uk/government/publications/healthy-child-programme-pregnancy-and-the-first-5-years-of-life

17. Physical - Monitoring Growth Part 2 [Internet]. [cited 2025 Oct 28]. Available from: https://resource.download.wjec.co.uk/vtc/2018-19/HSC18-19_4-1/_multi-lang/unit01/02-physical-monitoring-growth-part-2.html

18. Chadd K, Caute A, Pettican A, Enderby P. Operationalising routinely collected patient data in research to further the pursuit of social justice and health equity: a team-based scoping review. BMC Medical Research Methodology 2025 25:1 [Internet]. 2025 Jan 21 [cited 2025 Feb 18];25(1):1–21. Available from: https://bmcmedresmethodol.biomedcentral.com/articles/10.1186/s12874-025-02466-9

19. Harron K, Woodman J. Using administrative data to assess early-life policies. Lancet Public Health [Internet]. 2023 Jul 1 [cited 2025 Oct 21];8(7):e476–7. Available from: https://www.thelancet.com/action/showFullText?pii=S2468266723001275

20. Kane R, Wellings K, Free C, Goodrich J. Uses of routine data sets in the evaluation of health promotion interventions: opportunities and limitations. Health Educ [Internet]. 2000 Feb 1 [cited 2025 Oct 21];100(1):33–41. Available from: 10.1108/09654280010309030

21. Calanzani N, Vojt G, Weller D, Campbell C. Exploring challenges and strategies when using routine data in research: a systematic review [Internet]. Edinburgh University Press; 2017 [cited 2025 Oct 17]. Available from: https://researchonline.gcu.ac.uk/en/publications/exploring-challenges-and-strategies-when-using-routine-data-in-re

22. Bridges S, Henderson H, Ciesla K, Robinson K, Roberts K, Farrar D, et al. Born and Bred in (BaBi): an efficient, place-based birth e-cohort network. NIHR Open Research 2025 5:66 [Internet]. 2025 Aug 18 [cited 2025 Oct 17];5:66. Available from: https://openresearch.nihr.ac.uk/articles/5-66

23. Sohal K, Mason D, Birkinshaw J, West J, McEachan RRC, Elshehaly M, et al. Connected Bradford: a Whole System Data Linkage Accelerator. Wellcome Open Res [Internet]. 2022 [cited 2025 Oct 17];7:26. Available from: https://pmc.ncbi.nlm.nih.gov/articles/PMC9682213/

24. Henery P, Dundas R, Katikireddi SV, Leyland AH, Fenton L, Scott S, et al. A maternal and child health administrative cohort in Scotland: the utility of linked administrative data for understanding early years’ outcomes and inequalities. Int J Popul Data Sci [Internet]. 2024 [cited 2025 Sep 2];9(2):2402. Available from: https://pmc.ncbi.nlm.nih.gov/articles/PMC11977605/

25. Feng Q, Ireland G, Gilbert R, Harron K. Data Resource Profile: A national linked mother-baby cohort of health, education and social care data in England (ECHILD-MB). Int J Epidemiol [Internet]. 2024 Apr 11 [cited 2025 Oct 17];53(3). Available from: 10.1093/ije/dyae065

26. Peters M, Godfrey C, McInerney P, Munn Z, Tricco A, Khalil H. JBI Manual for Evidence Synthesis. 2024 [cited 2025 Oct 17]. Scoping reviews - JBI Manual for Evidence Synthesis. Available from: https://jbi-global-wiki.refined.site/space/MANUAL/355862497/10.+Scoping+reviews

27. Tricco AC, Lillie E, Zarin W, O’Brien KK, Colquhoun H, Levac D, et al. PRISMA extension for scoping reviews (PRISMA-ScR): Checklist and explanation. Ann Intern Med [Internet]. 2018 Oct 2 [cited 2024 May 15];169(7):467–73. Available from: https://www.acpjournals.org/doi/10.7326/M18-0850

28. Ayiku L, Levay P, Hudson T, Craven J, Barrett E, Finnegan A, et al. The medline UK filter: development and validation of a geographic search filter to retrieve research about the UK from OVID medline. Health Info Libr J [Internet]. 2017 Sep 1 [cited 2024 Feb 15];34(3):200–16. Available from: https://onlinelibrary.wiley.com/doi/full/10.1111/hir.12187

29. Stansfield C, Dickson K, Bangpan M. Exploring issues in the conduct of website searching and other online sources for systematic reviews: How can we be systematic? Syst Rev [Internet]. 2016 Nov 15 [cited 2025 Oct 17];5(1):1–9. Available from: https://systematicreviewsjournal.biomedcentral.com/articles/10.1186/s13643-016-0371-9

30. Owen-Jones E, Bekkers MJ, Butler CC, Cannings-John R, Channon S, Hood K, et al. The effectiveness and cost-effectiveness of the Family Nurse Partnership home visiting programme for first time teenage mothers in England: A protocol for the Building Blocks randomised controlled trial. BMC Pediatr [Internet]. 2013 Aug 6 [cited 2025 Oct 20];13(1):114. Available from: https://link.springer.com/articles/10.1186/1471-2431-13-114

31. Robling M, Lugg-Widger F, Cannings-John R, Sanders J, Angel L, Channon S, et al. The Family Nurse Partnership to reduce maltreatment and improve child health and development in young children: the BB:2–6 routine data-linkage follow-up to earlier RCT. Public Health Research. 2021 Feb;9(2):1–160.

32. Clarke JL, Ingram J, Johnson D, Thomson G, Trickey H, Dombrowski SU, et al. The ABA intervention for improving breastfeeding initiation and continuation: Feasibility study results. Matern Child Nutr [Internet]. 2020 Jan 1 [cited 2025 Oct 20];16(1):e12907. Available from: 10.1111/mcn.12907

33. MacArthur C, Jolly K, Ingram L, Freemantle N, Dennis CL, Hamburger R, et al. Antenatal peer support workers and initiation of breast feeding: cluster randomised controlled trial. BMJ [Internet]. 2009 Jan 30 [cited 2025 Oct 20];338(7691):392–5. Available from: https://www.bmj.com/content/338/bmj.b131

34. Relton C, Strong M, Renfrew MJ, Thomas K, Burrows J, Whelan B, et al. Cluster randomised controlled trial of a financial incentive for mothers to improve breast feeding in areas with low breastfeeding rates: the NOSH study protocol. BMJ Open [Internet]. 2016 Apr 1 [cited 2025 Oct 20];6(4):e010158. Available from: https://bmjopen.bmj.com/content/6/4/e010158

35. Sawtell M, Sweeney L, Wiggins M, Salisbury C, Eldridge S, Greenberg L, et al. Evaluation of community-level interventions to increase early initiation of antenatal care in pregnancy: Protocol for the Community REACH study, a cluster randomised controlled trial with integrated process and economic evaluations. Trials [Internet]. 2018 Mar 5 [cited 2025 Oct 20];19(1):1–13. Available from: https://link.springer.com/articles/10.1186/s13063-018-2526-6

36. Wiggins M, Sawtell M, Wiseman O, McCourt C, Greenberg L, Hunter R, et al. Testing the effectiveness of REACH Pregnancy Circles group antenatal care: Protocol for a randomised controlled pilot trial. Pilot Feasibility Stud [Internet]. 2018 Apr 25 [cited 2025 Oct 20];4(1):1–13. Available from: https://link.springer.com/articles/10.1186/s40814-018-0361-x

37. Wiggins M, Sawtell M, Wiseman O, McCourt C, Eldridge S, Hunter R, et al. Group antenatal care (Pregnancy Circles) for diverse and disadvantaged women: Study protocol for a randomised controlled trial with integral process and economic evaluations. BMC Health Serv Res [Internet]. 2020 Oct 7 [cited 2025 Oct 20];20(1):1– 14. Available from: https://link.springer.com/articles/10.1186/s12913-020-05751-z

38. Robling M, Cannings-John R, Lugg-Widger F. Using multiple routine data sources linked to a trial cohort to establish the longer-term effectiveness of specialist home visiting in England: main results of the BB:2-6 study of the Family Nurse Partnership. Int J Popul Data Sci [Internet]. 2022 [cited 2023 Nov 6]; Available from: https://ijpds.org/article/view/1829/3533

39. Robinson-Smith L, Menzies V, Cramman H, Wang Y, Fairhurst C, Hallett S, et al. EasyPeasy: Learning through play [Internet]. 2019. Available from: https://www.educationendowmentfoundation.org.uk

40. Vieira MC, Relph S, Copas A, Healey A, Coxon K, Alagna A, et al. The DESiGN trial (DEtection of Small for Gestational age Neonate), evaluating the effect of the Growth Assessment Protocol (GAP): Study protocol for a randomised controlled trial. Trials [Internet]. 2019 Mar 4 [cited 2025 Oct 21];20(1):1–14. Available from: https://link.springer.com/articles/10.1186/s13063-019-3242-6

41. Cavallaro F, Clery A, Gilbert R, van der Meulen J, Kendall S, Kennedy E, et al. Evaluating the real-world implementation of the Family Nurse Partnership in England: a data linkage study. Health and Social Care Delivery Research. 2024 May 1;12(11).

42. Lugg-Widger F V., Cannings-John R, Channon S, Fitzsimmons D, Hood K, Jones KH, et al. Assessing the medium-term impact of a home-visiting programme on child maltreatment in England: protocol for a routine data linkage study. BMJ Open [Internet]. 2017 Jul 1 [cited 2025 Oct 20];7(7):e015728. Available from: https://bmjopen.bmj.com/content/7/7/e015728

43. Mooney KE, Bywater T, Dickerson J, Richardson G, Hou B, Wright J, et al. Protocol for the effectiveness evaluation of an antenatal, universally offered, and remotely delivered parenting programme ‘Baby Steps’ on maternal outcomes: a Born in Bradford’s Better Start (BiBBS) study. BMC Public Health. 2023 Jan 28;23(1).

44. Tassie E, Langham J, Gurol-Urganci I, van der Meulen J, Howard LM, Pasupathy D, et al. An exploration of service use pattern changes and cost analysis following implementation of community perinatal mental health teams in pregnant women with a history of specialist mental healthcare in England: a national population-based cohort study. BMC Health Serv Res [Internet]. 2024 Dec 1 [cited 2025 Oct 20];24(1):1–10. Available from: https://link.springer.com/articles/10.1186/s12913-024-10553-8

45. Denison FC, Macgregor H, Stirrat LI, Stevenson K, Norman JE, Reynolds RM. Does attendance at a specialist antenatal clinic improve clinical outcomes in women with class III obesity compared with standard care? A retrospective case-note analysis. BMJ Open [Internet]. 2017 May 1 [cited 2025 Oct 20];7(5):e015218. Available from: https://bmjopen.bmj.com/content/7/5/e015218

46. Gurol-Urganci I, Waite L, Webster K, Jardine J, Carroll F, Dunn G, et al. Obstetric interventions and pregnancy outcomes during the COVID-19 pandemic in England: A nationwide cohort study. PLoS Med [Internet]. 2022 Jan 1 [cited 2025 Oct 20];19(1):e1003884. Available from: https://journals.plos.org/plosmedicine/article?id=10.1371/journal.pmed.1003884

47. Dundas R, Ouédraogo S, Gray R, Wood R, Bond L, Briggs A, et al. Assessment of the Health in Pregnancy Grant policy in Scotland: a natural experiment. The Lancet [Internet]. 2015 Nov 13 [cited 2025 May 27];386:S35. Available from: https://www.thelancet.com/action/showFullText?pii=S0140673615008739

48. Lugg-Widger F, Robling M, Lau M, Paranjothy S, Pell J, Sanders J, et al. Evaluation of the effectiveness of the Family Nurse Partnership home visiting programme in first time young mothers in Scotland: a protocol for a natural experiment. Int J Popul Data Sci [Internet]. 2020 Jan 30 [cited 2025 Oct 20];5(1):1154. Available from: https://pmc.ncbi.nlm.nih.gov/articles/PMC7473263/

49. Mooney KE, Bywater T, Hinde S, Richardson G, Wright J, Dickerson J, et al. A quasi-experimental effectiveness evaluation of the ‘Incredible Years Toddler’ parenting programme on children’s development aged 5: A study protocol. Lennox C, editor. PLoS One [Internet]. 2023 Sep 27 [cited 2023 Oct 3];18(9):e0291557. Available from: https://journals.plos.org/plosone/article?id=10.1371/journal.pone.0291557

50. Doi L, Morrison K, Astbury R, Eunson J, Horne MA, Jepson R, et al. Study protocol: a mixed-methods realist evaluation of the Universal Health Visiting Pathway in Scotland. BMJ Open [Internet]. 2020 Dec 1 [cited 2025 Oct 20];10(12):e042305. Available from: https://bmjopen.bmj.com/content/10/12/e042305

51. Harnett PH, Barlow J, Coe C, Newbold C, Dawe S. Assessing Capacity to Change in High-Risk Pregnant Women: A Pilot Study. Child Abuse Review [Internet]. 2018 Jan 1 [cited 2025 Oct 20];27(1):72–84. Available from: 10.1002/car.2491

52. Scourfield J, Webb CJR, Elliott M, Staniland L, Bywaters P. Are Child Welfare Intervention Rates Higher or Lower in Areas Targeted for Enhanced Early Years Services? Child Abuse Review [Internet]. 2021 Jul 1 [cited 2025 Oct 20];30(4):306–17. Available from: 10.1002/car.2696

53. Wood R, Wilson P. General practitioner provision of preventive child health care: Analysis of routine consultation data. BMC Fam Pract [Internet]. 2012 Aug 3 [cited 2025 Oct 20];13(1):1–8. Available from: https://link.springer.com/articles/10.1186/1471-2296-13-73

54. Leyland AH, Ouédraogo S, Nam J, Bond L, Briggs AH, Gray R, et al. Evaluation of Health in Pregnancy grants in Scotland: a natural experiment using routine data. Public Health Research. 2017 Oct;5(6):1–278.

55. Bennett L, Grant A, Jones S, Bowley M, Heathcote-Elliott C, Ford C, et al. Models for Access to Maternal Smoking cessation Support (MAMSS): A study protocol of a quasi-experiment to increase the engagement of pregnant women who smoke in NHS Stop Smoking Services. BMC Public Health. 2014 Oct 6;14(1).

56. Bauld L, Hackshaw L, Ferguson J, Coleman T, Taylor G, Salway R. Implementation of routine biochemical validation and an ‘opt out’ referral pathway for smoking cessation in pregnancy. Addiction [Internet]. 2012 Dec [cited 2025 Oct 20];107(SUPPL.2):53–60. Available from: 10.1111/j.1360-0443.2012.04086.x

57. Smith LK, Van Blankenstein E, Fox G, Seaton SE, Martínez-Jiménez M, Petrou S, et al. Effect of national guidance on survival for babies born at 22 weeks’ gestation in England and Wales: population based cohort study. BMJ Medicine [Internet]. 2023 Nov 7 [cited 2025 Oct 21];2(1):e000579. Available from: https://pmc.ncbi.nlm.nih.gov/articles/PMC10649719/

58. Mackay DF, Nelson SM, Haw SJ, Pell JP. Impact of Scotland’s Smoke-Free Legislation on Pregnancy Complications: Retrospective Cohort Study. PLoS Med [Internet]. 2012 Mar [cited 2025 Oct 20];9(3):e1001175. Available from: https://journals.plos.org/plosmedicine/article?id=10.1371/journal.pmed.1001175

59. Bell R, Glinianaia S V., Van Der Waal Z, Close A, Moloney E, Jones S, et al. Evaluation of a complex healthcare intervention to increase smoking cessation in pregnant women: interrupted time series analysis with economic evaluation. Tob Control [Internet]. 2018 Jan 1 [cited 2025 Oct 20];27(1):90–8. Available from: https://tobaccocontrol.bmj.com/content/27/1/90

60. Fair FJ, Soltani H. Association of child weight with attendance at a healthy lifestyle service among women with obesity during pregnancy. Matern Child Nutr [Internet]. 2024 Apr 1 [cited 2025 Oct 20];20(2):e13629. Available from: 10.1111/mcn.13629

61. Marmot M. Health equity in England: the Marmot review 10 years on. BMJ [Internet]. 2020 Feb 25 [cited 2021 Sep 24];368. Available from: https://www.bmj.com/content/368/bmj.m693

62. Mc Cord KA, Al-Shahi Salman R, Treweek S, Gardner H, Strech D, Whiteley W, et al. Routinely collected data for randomized trials: Promises, barriers, and implications. Trials [Internet]. 2018 Jan 11 [cited 2025 Oct 17];19(1):1–9. Available from: https://link.springer.com/articles/10.1186/s13063-017-2394-5

63. Foster V, Young A. The use of routinely collected patient data for research: A critical review. Health N Hav [Internet]. 2012 Jul [cited 2025 Oct 17];16(4):448–63. Available from: https://scholar.google.com/scholar_url?url= https://journals.sagepub.com/doi/pdf/10.1177/1363459311425513&hl=en&sa=T&oi=ucasa&ct=usl&ei=01HyaNK_BYePieoPxbqFoQM&scisig=AAZF9b-p8abIgDKcMS9VJtsKTKNI

64. GOV.UK. Early Years Foundation Stage Profile: 2020 Handbook. 2019 [cited 2024 Jan 10]; Available from: https://dera.ioe.ac.uk/id/eprint/34802/

65. Kendall S, Nash A, Braun A, Bastug G, Rougeaux E, Bedford H. Evaluating the use of a population measure of child development in the Healthy Child Programme Two Year Review. Policy Research Unit in the Health of Children, Young People and Families* Centre for Research in Primary and Community Care**. 2014;

66. Fraser C, Harron K, Barlow J, Bennett S, Woods G, Shand J, et al. Variation in health visiting contacts for children in England: cross-sectional analysis of the 2–2½ year review using administrative data (Community Services Dataset, CSDS). BMJ Open. 2022 Feb;12(2):e053884.

67. Henderson HC, Mooney KE, Morton K, Armour L, Lee D, Ahern S, et al. Use of the Ages and stages questionnaire in Bradford as a measure of Child Development (ABCD): a mixed methods study protocol. medRxiv [Internet]. 2025 Aug 16 [cited 2025 Sep 26];2025.08.14.25333745. Available from: https://www.medrxiv.org/content/10.1101/2025.08.14.25333745v1

