## Appendix 2 - Search terms for "Harnessing routinely collected data for the evaluation of early years interventions: insights from a scoping review of evaluation studies"

### Appendix 1: Search strategy

Table 2. Overview of search strategy and terms

| **Population** (COMBINE WITH OR)  “Early years”  Infant*  Infancy  Baby  Babies  Toddler*  Preschool  “First years”  “Early childhood”  Child*  Antenatal  Postnatal  Newborn*  Mother*  Parent*  Family  Families  Maternity  Maternal  Father  Paternal  Pregnancy  Pregnant  Perinatal  Paediatric*  Pediatric | **Concept 1** **(COMBINE WITH OR)**  Intervention  Programme  Support  Policy  Policies  “Randomised Control Trial”  RCT  TwiCS  “Trial within a Cohort Study”  QED  Quasi-experimental*  Difference-in-difference  “Regression discontinuity”  “Before and after”  “Case control”  “Time series”  “Instrumental variables”  “Propensity score matching” | **Concept 2** **(COMBINE WITH OR)**  Routine adj3 data  Routine adj3 outcome*  “Medical records”  “General practice data”  “GP data”  Administrative adj3 data  “Health event data”  “Electronic health records”  EHRs  “Matern* record*”  “Ages and Stages Questionnaire”  ASQ  “Early Years Foundation Stage Profile”  EYFSP  “Early Language Identification Measure”  ELIM  “Reception Baseline Assessment”  RBA | **Context**  NICE filter for screening UK studies, see (32)  for details. |
| --- | --- | --- | --- |

1. *truncates the word allowing multiple variations of the term.
2. Adj3 allows up to three words to be in between e.g. ‘routine’ and ‘data’.
3. Columns were combined with ‘and’
4. The default keyword search on databases on the Ovid platform is a multi-purpose search across several fields including title, abstract, original title, name of substance word, subject heading word, keyword heading word, protocol supplementary concept word, rare disease supplementary concept word, unique identifier.
