## Appendix 3 - Study data for "Harnessing routinely collected data for the evaluation of early years interventions: insights from a scoping review of evaluation studies"

| **Study reference** | **UK origin** | **Participants eligible for the intervention** | **Design and Sample size** | **Intervention description** | **Description of usage of routine data (outcome)** | **Outcome focus categorised** | **Data sources** | **Timepoint of measurement** |
| --- | --- | --- | --- | --- | --- | --- | --- | --- |
| **Study protocols** | | | | | | | | |
| Owen-Jones, E., Bekkers, M.J., Butler, C.C., Cannings-John, R., Channon, S., Hood, K., Gregory, J.W., Kemp, A., Kenkre, J., Martin, B.C. and Montgomery, A., 2013. The effectiveness and cost-effectiveness of the Family Nurse Partnership home visiting programme for first time teenage mothers in England: a protocol for the Building Blocks randomised controlled trial. BMC pediatrics, 13, pp.1-13. | England | Women aged 19 or under  Lives within the catchment area First pregnancy Recruited no later than 24 weeks gestation. Gillick competent to provide informed consent | Randomised control trial  Sample size N/A, protocol  * Aim to recruit a total of 1600 | The FNP is a structured, intensive programme during which participants receive up to 64 home based visits from a trained FNP family nurse from early pregnancy (of their first child) until the child is two years old. The aim of improving outcomes for the health, wellbeing, and social circumstances of young first-time mothers and their children. | Birth weight  Prenatal tobacco use  Emergency attendances and/or admissions within 2 years of birth  Second pregnancy within 2 years of first birth | See Owen-Jones (2013), is same study. | Maternity records, primary care records, and Hospital Episodes Statistics | Baseline, 34-36 weeks gestation, at birth, at 6, 12, 18 and 24 months following birth. |
| Bennett, L., Grant, A., Jones, S., Bowley, M., Heathcote-Elliott, C., Ford, C., Jones, A., Lewis, R., Munkley, M., Owen, C. and Petherick, A., 2014. Models for Access to Maternal Smoking cessation Support (MAMSS): a study protocol of a quasi-experiment to increase the engagement of pregnant women who smoke in NHS Stop Smoking Services. BMC Public Health, 14, pp.1-8. | Wales | Women who are pregnant and smoke or who have quit in the two weeks prior to their initial antenatal booking appointment | Quasi-experimental study  Sample size N/A, protocol | Referral to one of three models of service delivery where smoking cessation services are provided by 1) whole-time-equivalent (WTE) maternity support workers, 2) midwives or 3) smoking cessation advisors, all offering a flexible service designed to meet the needs of the women.  The core elements include: Strict adherence to NICE opt out smoking cessation pathway for pregnant women, including CO monitoring; Smoking cessation services being more closely aligned to maternity services (provided as part of the package of maternity care); Referral from midwife to smoking cessation support within 48 hours; Flexibility in service model with a women centred approach. | Primary: engagement with stop smoking service.  Secondary: pregnant smokers who set a quit date; pregnant smokers who quit at four weeks followup (CO verified); smoking status at the time of birth or third trimester and 52 weeks; and birth outcomes (low birth weight (<2500 g), preterm birth <37 weeks). | Smoking status | Health Board’s electronic Patient Administration System (PAS) and Maternity Information System | Antenatal appointments, face-to-face treatment sessions, and at 4-6 weeks after participants' quit dates |
| Relton, C., Strong, M., Renfrew, M.J., Thomas, K., Burrows, J., Whelan, B., Whitford, H.M., Scott, E., Fox-Rushby, J., Anoyke, N. and Sanghera, S., 2016. Cluster randomised controlled trial of a financial incentive for mothers to improve breast feeding in areas with low breastfeeding rates: the NOSH study protocol. BMJ open, 6(4), p.e010158. | Samples taken from 5 districts: Sheffield, North Derbyshire, Rotherham, Doncaster and Bassetlaw | All pregnant women aged 16+, resident in each ward, and with an estimated date of delivery between 18 February 2015 and 17 February 2016. | Cluster RCT  Sample size N/A, protocol | Nourishing Start for Health (NOSH) intervention - a structured financial incentive scheme to help increase breastfeeding rates in wards with low rates. | Breastfeeding rates from routine health records | Breastfeeding | district public health departments; local NHS trusts; Census Data; Hospital Episode Statistics | 6 to 8 weeks post birth |
| Lugg-Widger, F.V., Cannings-John, R., Channon, S., Fitzsimmons, D., Hood, K., Jones, K.H., Kemp, A., Kenkre, J., Longo, M., McEwan, K. and Moody, G., 2017. Assessing the medium-term impact of a home-visiting programme on child maltreatment in England: protocol for a routine data linkage study. BMJ open, 7(7), p.e015728. | England | Mothers and their child on the BB:0-2 trial and who were not withdrawn were recruited as nulliparous women aged 19 or under, living in one of 18 local authority FNP catchment areas; recruited by 24+6 weeks gestation, have conversational level of English and were able to consent to research | Observational study  Sample size N/A, protocol  *An estimated 1405 participants | A home-visiting approach with three overarching goals: to improve birth outcomes, optimise child health and development including reducing maltreatment, and promote economic self-sufficiency of mothers assessed the short-term impact of an intensive programme of antenatal and postnatal visiting by specially trained nurses to support young pregnant women in England. | primary outcome: CIN status at any point between birth and 6 years  Secondary outcomes: measures of maltreatment, intermediate FNP programme outcomes as well as child health, development and educational outcomes | Child abuse/neglect/maltreatment/protection  Child education/development  Child health | NHS Digital, Office for National Statistics and the Department for Education’s National Pupil Database | Up to 6 years |
| Sawtell, M., Sweeney, L., Wiggins, M., Salisbury, C., Eldridge, S., Greenberg, L., Hunter, R., Kaur, I., McCourt, C., Hatherall, B. and Findlay, G., 2018. Evaluation of community-level interventions to increase early initiation of antenatal care in pregnancy: protocol for the community REACH study, a cluster randomised controlled trial with integrated process and economic evaluations. Trials, 19, pp.1-13. | North and East London and Essex were targeted for study population | Women in the selected electoral wards who give birth, at a hospital enrolled in the study | Cluster RCT with process and economic evaluative  Sample size N/A protocol | A community-centred intervention that seeks to increase early initiation of antenatal care in communities | The primary outcome: proportion of pregnant women in each ward who have attended their antenatal booking appointment by the end of the 12th completed week of their pregnancy. Secondary outcomes: the proportion of women who have attended their antenatal booking appointment by 10 weeks and 0 days of pregnancy; antenatal admissions; emergency caesarean rates; gestation and weight at delivery; smoking at booking appointment and at birth; feeding method at discharge. | Service implementation  Birth outcome | NHS trust hospital data | Up to 10 weeks post birth |
| Vieira, M.C., Relph, S., Copas, A., Healey, A., Coxon, K., Alagna, A., Briley, A., Johnson, M., Lawlor, D.A., Lees, C. and Marlow, N., 2019. The DESiGN trial (DEtection of Small for Gestational age Neonate), evaluating the effect of the Growth Assessment Protocol (GAP): study protocol for a randomised controlled trial. Trials, 20, pp.1-14. | Majority of sites in London | Data from all women who give birth within an eligible maternity unit will be included within the study unless individuals specifically opt out of the study. Maternity units that have already fully implemented growth assessment protocols were not eligible. Data from women with multiple pregnancies or fetal congenital abnormalities will be excluded from the analysis. | Cluster RCT  Sample size N/A, protocol | Growth Assessment Protocol (GAP) is a programme developed by the Perinatal Institute that consists of the use of gestation-related optimal weight (GROW) customised charts, alongside a schedule of antenatal risk assessment for Small for Gestational Age (SGA), management protocols for suspected SGA foetuses, audit tools and training | Primary outcome: antenatal ultrasound detection of SGA (after 24 completed weeks of gestation) in infants who are also small for gestational age at birth.   Secondary neonatal outcomes: gestational age at birth, birthweight, head circumference, 5-min Apgar score < 7, Arterial cord pH < 7.1, Any respiratory support given at delivery, Length of stay at each neonatal level of care, major and minor neonatal morbidity, Antepartum or intrapartum stillbirth, Neonatal death (early or late), Death before neonatal discharge (after 28 days of birth), and Cause of death  Secondary maternal outcomes: length of stay in hospital, Induction of labour, Mode of delivery (including caesarean section rates), Postpartum haemorrhage, Rates of 3rd or 4th degree perineal tear, Length of stay in hospital, and Breastfeeding at discharge.  Protocol says most data will be acquired from routine hospital systems, not specified which. | Child health | (i) maternal/perinatal characteristics and outcomes (maternity EPR), (ii) characteristics and outcomes of infants admitted to neonatal care (neonatal EPR), (iii) timing and findings of antenatal ultrasound scans (ultrasound EPR) and (iv) hospital activity, e.g. number of antenatal clinic visits (hospital administrative EPR). | At birth |
| Wiggins, M., Sawtell, M., Wiseman, O., McCourt, C., Greenberg, L., Hunter, R., Eldridge, S., Haora, P., Kaur, I. and Harden, A., 2018. Testing the effectiveness of REACH Pregnancy Circles group antenatal care: protocol for a randomised controlled pilot trial. Pilot and Feasibility Studies, 4, pp.1-13. | 3 London boroughs | Women who are currently pregnant and registering (or registered) for antenatal care at the participating NHS Trust maternity service and live within the working areas of the local midwife group facilitators and have an estimated delivery date that fits with those of a proposed group. | Pilot RCT with process and economic evaluative  Sample size N/A, protocol | Antenatal care model 'Pregnancy Circles' - consist of 8-12 PW with estimated delivery dates in same 2-4 week period. Aim is to empower women, give them a 'voice'. enhance decision making and ultimately tailor antenatal care more closely to their own needs. aim to increase sense of control around birthing and therfore better birthing experience, particularly in women from ethnically, culturally and linguistically diverse and disadvantaged areas who are more likely to experience worse outcomes | Spontaneous vaginal birth (primary outcome)   Attendance at ANC (number of regular appointments attended)   Caesarean delivery (planned, emergency, none)   Epidural/spinal analgesia use in labour   Infant birthweight, defined as low if less than 2500 g   Gestational age at delivery, dichotomized as term or preterm (less than 37 weeks)   Breastfeeding initiation | Service implementation  Birth outcome | Maternity records | At birth |
| Doi, L., Morrison, K., Astbury, R., Eunson, J., Horne, M.A., Jepson, R., Marryat, L., Ormston, R. and Wood, R., 2020. Study protocol: a mixed-methods realist evaluation of the Universal Health Visiting Pathway in Scotland. BMJ open, 10(12), p.e042305. | Scotland | Not specified, but parents in receipt of health visiting in routine data | Mixed-methods realist evaluative  Sample size N/A, protocol  *80 HVs, 60 parents. 75 case notes. | The Universal Health Visiting Pathway (UHVP) was introduced in Scotland in 2015. The UHVP refocuses the role of the health visitor and includes changes to caseload weighting and management; intervention delivery; education, training and resources; and visiting patterns. | Parental smoking  Developmental concerns  Child BMI  Child protection interventions | Smoking status  Child abuse/ neglect/ maltreatment/protection,  Child education/ development,  Child BMI/weight | Routine data captured via health or social work records, accessed through NHS Public Health Scotland. | Various time points for both child and parents until child is 3 years |
| Lugg-Widger, F., Robling, M., Lau, M., Paranjothy, S., Pell, J., Sanders, J., White, J. and Cannings-John, R., 2020. Evaluation of the effectiveness of the Family Nurse Partnership home visiting programme in first time young mothers in Scotland: a protocol for a natural experiment. International Journal of Population Data Science, 5(1). | Scotland (10 health boards nationally) | 1. Living in an FNP-recruiting NHS Health Board  2. First time mothers-to-be (women are eligible if a previous pregnancy resulted in a miscarriage, stillbirth or termination)  3. Aged 19 years or younger at time of last menstrual period (LMP)  4. Less than 28+6 weeks gestation at enrolment into FNP | Observational, natural experiment  Sample size N/A, protocol | The Nurse Family Partnership (NFP) is an intensive preventative home-visiting service which involves up to 64 structured home visits across pregnancy, infancy and toddlerhood by specially recruited and trained family nurses from early pregnancy until children are 2 years of age. The programme aims to address the problems of poor birth outcomes, child abuse and neglect, and the diminished economic self-sufficiency of mothers. | Maternal characteristics and information and child health and educational outcomes. | Child education/ development  Child health | NHS National Services Scotland (NSS) FNP Scottish Information System (FNP SIS) National Records for Scotland (NRS) | From conception to child is 2 and medium outcomes, and during first year of childs school |
| Wiggins, M., Sawtell, M., Wiseman, O., McCourt, C., Eldridge, S., Hunter, R., Bordea, E., Mustard, C., Hanafiah, A., Hatherall, B. and Holmes, V., 2020. Group antenatal care (Pregnancy Circles) for diverse and disadvantaged women: study protocol for a randomised controlled trial with integral process and economic evaluations. BMC Health Services Research, 20, pp.1-14. | England | Women who are 16 years old and over, are part of the cohort of women cared for by the team delivering the intervention, have an estimated delivery date that fits with those of a proposed group and do not have a documented learning disability. | Parallel group RCT  Sample size N/A, protocol | The Pregnancy Circle model will consist of eight antenatal group sessions, reflecting the standard schedule for antenatal care in the UK, each of which will last two hours. The first Pregnancy Circle session will take place at approximately 16 weeks of pregnancy. Subsequent sessions will adhere to the standard NHS antenatal care schedule for primigravida women. | The primary outcome is a ‘healthy baby’ composite.  Secondary outcomes are health and maternity related. | Child health  Birth outcome | NHS trust electronic patient record data, supplemented where there are gaps via access to paper maternity notes. | At birth (secondary outcomes) and at 1 month postnatal (primary outcome) |
| Mooney, K.E., Bywater, T., Dickerson, J., Richardson, G., Hou, B., Wright, J. and Blower, S., 2023. Protocol for the effectiveness evaluation of an antenatal, universally offered, and remotely delivered parenting programme ‘Baby Steps’ on maternal outcomes: a Born in Bradford’s Better Start (BiBBS) study. BMC Public Health, 23(1), p.190. | Bradford | 1. Currently pregnant and have not exceeded 24 weeks gestation 2. Lives in the Better Start Bradford area. 3. Have consented to a referral to Baby Steps. | Quasi-experimental  Sample size N/A, protocol | Baby Steps, a relationship-based postnatal and antenatal parent education programme for 'vulnerable and socially excluded parents who often face challenges and 'overload' in pregnancy and early parenting | Data linked to routinely collected birth outcome (descriptive differences across delivery method, gestation period, and baby weight) by maternity services and Health Visitors | Maternal mental health  Maternal healthcare use,  Birth outcome | Maternity services and health visitors | Through pregnancy and up to one year post birth. |
| Mooney, K.E., Bywater, T., Hinde, S., Richardson, G., Wright, J., Dickerson, J. and Blower, S.L., 2023. A quasi-experimental effectiveness evaluation of the’Incredible Years Toddler’parenting programme on children’s development aged 5: A study protocol. Plos one, 18(9), p.e0291557. | Bradford | Participants must be enrolled in the BiBBS cohort.   Eligibility criteria for the intervention group are: Consented to participate in BiBBS Parent consented to a referral and enrolled in IY-T | Quasi-experimental  Sample size N/A, protocol | The Incredible Years (IY) parent programmes (www.incredibleyears.com), which are parent education and training group-based interventions informed by social learning theory and designed to enhance the social and emotional wellbeing of children aged 0–12 years. These manualised programmes are typically delivered by trained facilitators to groups of 10–12 parents for two hours a week for 12–20 weeks. This study evaluates Incredible Years (Toddler). | For the primary outcome: child social and emotional development.  For the secondary outcome: child early years development | Child education/ development | Routine education records: Early Years Foundation Stage Profile Profile (EYFSP) | Child age 5 years old |
| **Studies with results** | | | | | | | | |
| MacArthur, C., Jolly, K., Ingram, L., Freemantle, N., Dennis, C.L., Hamburger, R., Brown, J., Chambers, J. and Khan, K., 2009. Antenatal peer support workers and initiation of breast feeding: cluster randomised controlled trial. Bmj, 338. | Deprived urban area of Birmingham | Every pregnant women registered in the trust | Cluster RCT  2511 (2398 had outcome data) | Community based antenatal breastfeeding service using peer support workers | Primary outcome was initiation of breastfeeding | Breastfeeding | Hospital maternity records | At the time of delivery or by the time of hospital discharge |
| Bauld, L., Hackshaw, L., Ferguson, J., Coleman, T., Taylor, G. and Salway, R., 2012. Implementation of routine biochemical validation and an ‘opt out’referral pathway for smoking cessation in pregnancy. Addiction, 107, pp.53-60. | Dudley and South Birmingham | If a woman was a smoker, or had recently quit smoking | Observational pilot  6075 antenatal bookings total | Introduction of a referral pathway where women 'opt out' of passing their details to local smoking cessation services (rather than 'opt in'), after being identfied as a smoker via direct question or CO test. | Acceptance of opt out referral to the local stop smoking service, measured levels of CO, cotinine levels collected through routine urine sampling and follow-up of referred women by the local stop smoking service. | Smoking status | Maternity booking data | 12-week maternity scan |
| Robling, M., Lugg-Widger, F., Cannings-John, R., Sanders, J., Angel, L., Channon, S., Fitzsimmons, D., Hood, K., Kenkre, J., Moody, G. and Owen-Jones, E., 2021. The Family Nurse Partnership to reduce maltreatment and improve child health and development in young children: the BB: 2 6 routine data-linkage follow-up to earlier RCT. Public Health Research, 9. | England | See Owen-Jones (2013), is same study. | RCT  Data for 1537 mothers and 1547 children | See Owen-Jones (2013), is same study. | Recording of child-in-need status (i.e. child is unlikely to achieve or maintain a reasonable level of health or development, or whose health and development is likely to be significantly or further impaired without provision of services, or a child who is disabled).  Additional objective measures of maltreatment: referral to social services (overall, child protection referral, child-in-need referral), child protection registration, child-in-need categorisation, looked-after status (mother, child)  Associated measures of maltreatment: recorded injuries and ingestions, non-attendance rates for hospital appointments  Maternal outcomes: subsequent pregnancies Child health and developmental and educational outcomes: special educational needs, early educational attendance and assessments (Early Years Foundation Stage profile, Key Stage 1). | Child abuse/ neglect /maltreatment/protection  Child education/ development  Maternal reproductivity | Hospital Episode Statistics data (NHS Digital), social care and educational data (National  Pupil Database) and abortions data (Department of Health and Social Care). | Up to 7 years post birth (based on KS1) |
| Mackay, D.F., Nelson, S.M., Haw, S.J. and Pell, J.P., 2012. Impact of Scotland's smoke-free legislation on pregnancy complications: retrospective cohort study. PLoS medicine, 9(3), p.e1001175. | Scotland | Infants delivered in Scotland between 1 January 1996 and 31 December 2009. | Observational, retrospective cohort  716,941 | The Smoking, Health and Social Care (Scotland) Bill prohibited smoking in all enclosed public places and workplaces from 26 March 2006, with the primary aim of reducing exposure to environmental tobacco smoke (ETS) in public places. | The primary outcomes: preterm delivery and small for gestational age.  Secondary outcomes included low birth weight; spontaneous preterm labour; preterm delivery; and very small for gestational age. | Child health,  Birth outcome | The Scottish Morbidity Record (SMR2) | At birth |
| Wood, R. and Wilson, P., 2012. General practitioner provision of preventive child health care: analysis of routine consultation data. BMC Family Practice, 13, pp.1-8. | 30 GP Practices in Scotland | Pre-school children aged 0-4 | Observational, before and after  11,214 total | Change in policy/recommendations reduced number of universal child health surveillance reviews for pre-schoolers. This study aimed to quantify GPs’ provision of different types of preventive care to pre-school children before and after the changes to the CHS system. | Child health review by GP | Service implementation | GP consultation records | 6-8 weeks. 8–9, 21–24, 39–42, and 48 months. |
| Gale, C., Santhakumaran, S., Nagarajan, S., Statnikov, Y. and Modi, N., 2012. Impact of managed clinical networks on NHS specialist neonatal services in England: population based study. Bmj, 344. | England, Wales and Northern Ireland | Babies born alive at 27-28 weeks gestation | Observational, before and after  3522 before, 2919 after | Reorganisation of the neonatal services into managed clinical networks, in which clusters of hospitals providing different levels of specialist care work in collaboration | (1) Proportion of babies born at 27-28 weeks’ gestation at a hospital providing the highest specialist neonatal intensive care activity, (2) proportion transferred within the first 24 hours after birth (acute transfer), (3) proportion transferred between 24 hours and 28 days after birth (late transfer), and (4) proportion of babies in twin or higher order birth sets who are separated by transfer, (5) rate of transfers in singleton and multiple births, (6) changes in 28 day mortality | Birth outcome  Child health | National Neonatal Research Database held by the Neonatal Data Analysis Unit.  Encrypted maternal NHS numbers were used to link multiple birth sets. | 28 days post birth |
| Denison, F.C., MacGregor, H., Stirrat, L.I., Stevenson, K., Norman, J.E. and Reynolds, R.M., 2017. Does attendance at a specialist antenatal clinic improve clinical outcomes in women with class III obesity compared with standard care? A retrospective case-note analysis. BMJ open, 7(5), p.e015218. | Two hospitals, Edinburgh | Singleton pregnancy, who gave birth between 2008 December 2014, must have been booked onto the specialist appoitnments no later than 20 weeks gestation, in the Lothian area, with class III obesity | Observational cohort  Specialised obesity clinic (n=511) compared with standard antenatal care (n=502) | A specialist antenatal clinic for women with class III obesity with the aim of improving maternal and offspring outcomes, such as hypertension, diabetes, delivery method and blood loss at delivery and antenatal obstetric triage attendedces. | Maternal outomes: hypertension, diabetes, onset of labour, delivery method, blood loss at delivery and antenatal obstetric triage attendances.   Child outcomes: gender, birth weight, birthweight centile, macrosomia , low birth weight, gestation of delivery, preterm birth and outcome. | Birth outcome  Maternal health | Maternity electronic patient records database TRAK (supplied by Intersystems), clinical biochemistry database APEX (ApexHealthware) and the neonatal unit electronic patient records database BadgerNet (supplied by Clevermed) systems | Midwife appointments and at birth |
| Harnett, P.H., Barlow, J., Coe, C., Newbold, C. and Dawe, S., 2018. Assessing capacity to change in high‐risk pregnant women: a pilot study. Child abuse review, 27(1), pp.72-84. | Oxfordshire | Women had at least one high-risk criteria, and were between 18 and 28 weeks pregnant. | Quasi-experimental  Pre-birth assessment and care pathway (n = 35), routine care (n = 33) | Pre-birth assessment and care pathway based on the capacity to change model. The Parents Under Pressure (PuP) programme was developed for high-risk families with potential or current involvement in child protection services. | Standardised measures were completed at referral and post-birth (2 months). The child protection status of the infants was obtained from administrative records at birth and at 12 months | Child abuse/ neglect/ maltreatment/protection | Maternal health records & child protection status via administrative records | Maternal: entry to pathway and 2 months postnatal.   Child: birth and at 12 months |
| Leyland, A.H., Ouédraogo, S., Nam, J., Bond, L., Briggs, A.H., Gray, R., Wood, R. and Dundas, R., 2017. Evaluation of health in pregnancy grants in Scotland: a natural experiment using routine data. Public Health Research, 5(6). | Scotland | Women in Great Britain and Northern Ireland reaching 25 weeks of pregnancy if they had sought health advice from a doctor or midwife | Observational, natural experiment  525,400 singleton births | Health in Pregnancy grant (a universal, unconditional cash transfer of £190 for women in Great Britain and Northern Ireland reaching 25 weeks of pregnancy if they had sought health advice from a doctor or midwife) | Birthweight was the primary outcome.  Secondary outcomes were maternal behaviour [gestation at booking (i.e. the first antenatal appointment with a health-care professional), booking before 25 weeks and maternal smoking during this pregnancy], measures of size (birthweight), crown-to-heel length and head circumference], measures of stage (gestational age), weight for dates (standardised, small and large for gestational age)] and birth outcomes (elective caesarean section, emergency caesarean section, stillbirths, neonatal deaths and 5-minute Apgar score). | Birth outcome  Smoking status | Maternal hospital discharge forms linked to the birth registration system via Scottish maternity and neonatal database held by the Information and Services Division at the NHS National Services Scotland. | Post birth |
| Bell, R., Glinianaia, S.V., van der Waal, Z., Close, A., Moloney, E., Jones, S., Araújo-Soares, V., Hamilton, S., Milne, E.M., Shucksmith, J. and Vale, L., 2018. Evaluation of a complex healthcare intervention to increase smoking cessation in pregnant women: interrupted time series analysis with economic evaluation. Tobacco Control, 27(1), pp.90-98. | North East England | All pregnant women | Quasi-experimental, interrupted time series  37,726 births | BabyClear: skills training for healthcare and smoking cessation staff; universal carbon monoxide monitoring with routine opt-out referral for smoking cessation support; provision of carbon monoxide monitors and supporting materials; and an explicit referral pathway and follow-up protocol. | Smoking status at delivery, quitting of smoking during pregnancy, and referral for smoking cessation advice. | Smoking status | Electronic maternity records from hospitals & referral, appointment, and quit attempt/status data from smoking cessation services  Data were linked using NHS numbers or mothers' DOBs and postcodes | Pregnancy and at delivery.   Data from smoking cessation services included referral dates, appointments, quit dates and quit status at 4 weeks. |
| Clarke, J.L., Ingram, J., Johnson, D., Thomson, G., Trickey, H., Dombrowski, S.U., Sitch, A., Dykes, F., Feltham, M., MacArthur, C. and Roberts, T., 2020. The ABA intervention for improving breastfeeding initiation and continuation: feasibility study results. Maternal & child nutrition, 16(1), p.e12907. | England | Women aged 16 years or older and pregnant with their first child. | RCT  103 | The 'Assets-based feeding help Before and After birth (ABA) intervention' is designed to be inclusive and improve infant feeding behaviours and breastfeeding rates. | Routinely collected health visitor data: breastfeeding outcomes. | Breastfeeding | Health visiting | 8 weeks post birth |
| Scourfield, J., Webb, C.J., Elliott, M., Staniland, L. and Bywaters, P., 2021. Are child welfare intervention rates higher or lower in areas targeted for enhanced early years services?. Child Abuse Review, 30(4), pp.306-317. | Wales | Data Eligibility: a list of areas eligible for Flying Start Services; an anonymised individual-level dataset of looked after children; anonymised individual-level data on children on the child protection register. | Cross-sectional area level study, quasi-experimental  1784 out of a Welsh total of 1909 LSOAs, including 467 out of 469 LSOAs where the Flying Start intervention operate. | Flying Start: aiming to improve life opportunities and outcomes for children in the most deprived areas of Wales. Comprised of 4 key elements: free, high-quality, part-time childcare for 2-to-3-year-olds; enhanced health visiting services, with one health visitor per 110 children; access to parenting support; and support for children's speech, language and communication skills. | Child welfare rates: calculated as number of children in care or on child protection registers (i.e. these two numbers combined) per 10,000 child population. | Child abuse/ neglect/ maltreatment/protection | 1. Welsh Government list of LSOAs and using local intelligence about areas of need. 2. 2015 Census Data 3. 22 Local Authorities child protection register 2015. | Up to child age 3 |
| Hildersley, R., Easter, A., Bakolis, I., Carson, L. and Howard, L.M., 2022. Changes in the identification and management of mental health and domestic abuse among pregnant women during the COVID-19 lockdown: regression discontinuity study. BJPsych open, 8(4), p.e96. | Two south london NHS hospitals | From time 1 October 2018 to 29 August 2020 from maternity services in two south london NHS hospitals. Each pregnancy must be recorded using eLIXIR database. | Quasi-experimental, regression discontinuity  26441 (24975 had completed data) | The introduction of the COVID-19 lockdown policy on 23 March 2020 acts as the first intervention of interest, followed by the start of lifting of the COVID-19 lockdown policy on 10 May 2020. | Identification of domestic violence and depressive symptoms | Maternal mental health  Domestic violence | Maternity and mental health records held within the Early Life Cross-Linkage in Research (eLIXIR) Partnership database | During pregnancy |
| Gurol-Urganci, I., Waite, L., Webster, K., Jardine, J., Carroll, F., Dunn, G., Frémeaux, A., Harris, T., Hawdon, J., Muller, P. and van der Meulen, J., 2022. Obstetric interventions and pregnancy outcomes during the COVID-19 pandemic in England: a nationwide cohort study. PLoS medicine, 19(1), p.e1003884. | England | All women with a record in HES of a singleton birth during the pandemic period (23 March 2020 to 22 February 2021) and the pre-pandemic period (the corresponding calendar period 1 year earlier: 23 March 2019 to 22 February 2020) | Observational, nationwide cohort  948,020 | Changes in maternity care due to the Covid-19 pandemic. | Stillbirth (fetal death at ≥24 weeks’ gestation), preterm birth (less than 37 weeks’ gestation), small for gestational age (SGA) at birth (defined as birthweight < 10th centile using population-based centile charts [18]), induction of labour, mode of birth (instrumental vaginal birth, elective cesarean section, emergency cesarean section, or unassisted vaginal birth), prolonged maternal length of stay (3 or more days after birth), and maternal readmission within 42 days of birth | Birth outcome | Hospital episode Statistics | Birth to 42 days post birth |
| Dundas, R., Boroujerdi, M., Browne, S., Deidda, M., Bradshaw, P., Craig, P., McIntosh, E., Parkes, A., Wight, D., Wright, C. and Leyland, A.H., 2023. Evaluation of the Healthy Start voucher scheme on maternal vitamin use and child breastfeeding: a natural experiment using data linkage. Public Health Research, 11(11), pp.1-101. | Scotland | Pregnant women and families with children under 4 years of age with low incomes or in receipt of certain means-tested benefits, all pregnant women aged < 18 years and all mothers aged < 18 years | Natural experiment, Growing Up in Scotland cohort  (n = 2240), respondents to the 2015 Infant Feeding Study (n = 8067) | The Healthy Start voucher, a means-tested scheme that provides vouchers worth £3.10 per week to spend on liquid milk, formula milk, fruit and vegetables. | Birthweight, low birthweight, preterm birth, gestational age and morbidity in first year/up to 3 years. | Birth outcome | Routinely collected NHS data: birth records, hospital records, deaths data, A&E attendance, SIRS Immunisation records, CHSP Health Visitor, MIDAS dental inspection. | Up to 3 years post birth for morbidity, other outcomes in between |
| Smith, L.K., van Blankenstein, E., Fox, G., Seaton, S.E., Martínez-Jiménez, M., Petrou, S., Battersby, C. and MBRRACE-UK Perinatal Surveillance Group, 2023. Effect of national guidance on survival for babies born at 22 weeks’ gestation in England and Wales: population based cohort study. BMJ medicine, 2(1), p.e000579. | England and Wales | Babies born (and whose mothers were resident) in England and Wales between 1 January 2018 and 31 December 2021 (inclusive) at 22+0 to 22+6 weeks’ gestational age. | Observational, cohort  5623 | The British Association of Perinatal Medicine developed a new consensus based framework for practice,11 updating guidelines from 2008. This new framework recommends that survival focused care may include babies from 22 weeks’ gestation following assessment of risk and discussion with parents. | Baby survivial rates: survival to admission for neonatal care, length of neonatal unit stay in days, survival to discharge from neonatal care (discharge home or to other healthcare settings, such as paediatric ward or intensive care unit), and survival to discharge without major morbidity. | Birth outcome  Child health | (1) mothers and babies: reducing risk through audits and confidential enquiries across the UK (MBRRACE-UK) and (2) the national neonatal research database (NNRD). | Babies born at 22 weeks gestation in neonatal units |
| Robinson-Smith, L., Merrell, C., Menzies, V., Cramman, H., Fairhurst, C., Hugill-Jones, J., Wang, Y., Hallett, S.,  Beckmann, N., Torgerson, C., & Stothard, S.(2019). EasyPeasy: Learning through play. Addendum Report. Education Endownment Foundation. <https://d2tic4wvo1iusb.cloudfront.net/production/documents/projects/EasyPeasy-Addendum-Report-Final.pdf?v=1748272886> | England | Children: children aged three to four years in school nurseries who were moving into the Reception year at school in the September following the intervention.  Nurseries: state-funded primary schools in England; with three-year-old pupils in their school population who had not previously been involved with EasyPeasy; and had an average ever-Free School Meal (FSM) percentage of >30% overall. | Cluster RCT  1488 pupils. 1205 pupils randomised, 1203 followed up using routine data | EasyPeasy provides game ideas to the parents of preschool children to encourage play-based learning at home, with the aim of developing children’s language development and self-regulation. | Communication and Language (CL) EYFSP score | Child education/ development | National Pupil Database data | immediately after intervention |
| Cavallaro, F., Clery, A., Gilbert, R., van der Meulen, J., Kendall, S., Kennedy, E., Phillips, C. and Harron, K., 2024. Evaluating the real-world implementation of the Family Nurse Partnership in England: a data linkage study. Health and social care delivery research, 12(11), pp.1-223. | England (136 local authorities) | Mothers aged 13–19 at last menstrual period (LMP) with their first live birth between April 2010 and March 2019, living in a FNP catchment area and their firstborn child(ren). | Observational, quasi-experimental  - Total mothers (n = 59,010) - Mothers enrolled in FNP (n = 29,505) - Mothers never enrolled in FNP (n = 29,505) | Family Nurse Partnershio (FNP) is an intensive nurse home visiting programme for first-time adolescent mothers. It includes up to 64 home visits by a family nurse from early pregnancy until the child's second birthday. | Main Outcomes (3 Outcome Domains, 13 Child Outcomes, 5 Maternal Outcomes):  1) Indicators of child maltreatment up to age 7 2) Child health and development outcomes (healthcare use up to age 7 and education between ages 5-7) 3) Maternal hospital utilisation (up to 7 years following delivery) and educational outcomes (up to 2 years following delivery) | Child education/ development  Child healthcare use Maternal healthcare use  Maternal education  Maternal reproductivity  Child abuse/ neglect/ maltreatment | Hospital Episode Statistics (Hospital Admissions Data) National Pupil Database (Education and Social Care Data) Family Nurse Partnership Information System | Between birth and up to 7 years after birth, depending on the outcome |
| Fair, F.J. and Soltani, H., 2024. Association of child weight with attendance at a healthy lifestyle service among women with obesity during pregnancy. Maternal & Child Nutrition, 20(2), p.e13629. | Two sites in England | Pregnant women with a BMI ≥ 35 kg/m2. | Observational, cohort  1301 mothers | Healthy Lifestyle service: a low‐intensity intervention to pregnant women with a BMI ≥ 35 kg/m2 at their first antenatal appointment which incorporated a visit at 16 weeks of gestation, with additional follow‐up visits available if the woman wanted them. The aim of the clinic was to encourage and support women to make positive lifestyle choices and behavioural changes during pregnancy to optimise GWG and improve birth outcomes. | Attendance at the antenatal healthy lifestyle service, maternal sociodemographic data and GWG & pregnancy data.  Weight of child. | Child BMI/weight  Birth outcome | Maternal: hospital records  Child: National Child Measurement Programme & SystmOne | Children: 6–8 weeks, 9–12 months, 4–5.5 years |
| Tassie, E., Langham, J., Gurol-Urganci, I., van der Meulen, J., Howard, L.M., Pasupathy, D., Sharp, H., Davey, A., O’Mahen, H., Heslin, M. and Byford, S., 2024. An exploration of service use pattern changes and cost analysis following implementation of community perinatal mental health teams in pregnant women with a history of specialist mental healthcare in England: a national population-based cohort study. BMC health services research, 24(1), p.359. | England | All women residing and giving birth in England with an onset of pregnancy on or after the 1st April 2016 and who gave birth on or before 31st March 2018 with pre-existing mental illness (defined as those with a previous secondary mental health care contact) | Observational, cohort  70,323 (complete data for 70,082) | Access to a Community Perinatal Mental Health Team (CPMHT) during the perinatal period | Service use pattern and cost changes | Service implementation, Maternal mental health | Mental Health Services Dataset (MHSDS), Hospital Episode Statistics (HES), and Personal Demographic Service (PDS) Birth Notification Data | N/A, service level. |

### Strengths and limitations noted in study discussion sections

| **Shortened reference** | **Strengths and limitations noted regarding routine data** |
| --- | --- |
| Vieira et al., (2019) | None |
| Owen-Jones et al., (2013) | None |
| Robling et al., (2021) | Limitations: • Routine data limits the scope of what outcomes can be examined, and the results are reliant on the quality of data coding at source.  • Routine data may lack the measurement properties that research instruments can offer (e.g. the EYFSP). |
| Bennett et al., (2014) | None |
| Relton et al., (2016) | Limitations: • concern in accuracy of routine data, as infant feeding reports rely on information exchanged between mother and her healthcare provider |
| Lugg-Widger et al., (2017) | Strengths • reduction in cost and participant burden • minimisation of bias in self-reporting • potential to follow up over time, e.g. criminal justice and welfare data |
| Sawtell et al., (2018) | Strength: • limited missing data can be expected |
| Wiggins et al., (2018) | None |
| Doi et al., (2020) | Limitations: • Only data included in routine data collections can be used which limits the outcomes included. |
| Lugg-Widger et al., (2020) | Limitations: • limited to outcomes available from routinely collected data.  • women who were approached but not enrolled to the programme were removed from the Cases, but those who might have been approached and subsequently not enrolled will still be present in the control group. Since access to these individuals’ data was not permitted, women of similar characteristics in the Controls cannot be identified to remove them. W • setting up the agreements took time and posed a risk to the delivery of the project |
| Wiggins et al., (2020) | None |
| Mooney et al., (2023) | None |
| Mooney et al., (2023) | Limitations: • There is a paucity of routinely collected data within the early years of a child’s life, which limited our choice of an outcome measure. Whilst the EYFSP is a well-timed measure and represents a construct of interest to this study, other measures relating to outcomes that IY-T aims to effect would have been beneficial (e.g. of parent-child bonding) [56].  • As collection of EYFSP data was paused during the COVID pandemic, there will inevitably be missing outcome data for a small number of children.   Strengths: • However, it is worth noting that EYFSP and other routinely collected health and education outcome data is regularly used as the basis for most policy-making decisions and this study reflects real-world conditions. |
| MacArthur et al., (2009) | Limitations: • data on initiation of breast feeding were obtained from the routinely collected maternity records, which are not generally considered to be as error free as data specifically collected by a research team.   Strengths: • this allowed a low loss to follow-up, at only 5%, and the quality of the data was similar across trial groups.  • primary care trusts in the UK use such hospital based data to assess their targets for initiation of breast feeding |
| Bauld et al., (2012) | Limitations:  • local services collecting limited routine data |
| Mackay et al., (2012) | Strengths: • routinely collected data are subjected to regular quality assurance checks, and their quality is high.  Limitations: • no access to reliable data on maternal obesity and height |
| Wood and Wilson (2012) | Limitations: • lack of data on care apart from routine child health reviews provided by Health Visitors means that the overall impact of the changes to the CHS system on the amount, content, and distribution of HV care (and how this relates to changes in GP provision of preventive care) therefore cannot be directly assessed |
| Gale et al., (2012) | Limitation: • only a single source of aggregated survival data was available before reorganisation  • could not fully account for clustering by hospital or area in epoch one due to data availability |
| Denison et al., (2017) | Strengths: • results are relevant to clinical practice  • there was a relatively low proportion of missing data. |
| Harnett et al., (2017) | Limitations: • it was not possible to obtain information on the nine closed cases in routine care as administrative data were not available • routine care does not contain standardised measures |
| Leyland et al., (2017) | Limitations: • Ethnicity was poorly recorded in the routine data set.  Strengths: • Increased power through use of routine data meant small changes with no clinical relevance could be detected. |
| Bell et al., (2018) | Limitations: • organisations collect different variables and define them differently • high levels of missing data. |
| Clarke et al., (2020) | Strengths: • UK collection of routine data for feeding method at 8 weeks by health visitors facilitates high completion for the primary outcome |
| Scourfield et al., (2021) | Limitation: • data for child protection registration and children looked after combined come from just 20 Welsh local authorities, due to substantial amount of missing LSOA data on children looked after from two local authorities |
| Hildersley et al., (2022) | Limitations: • Despite the large sample size, lack of power due to low rates of DVA recorded within maternity settings. |
| Gurol-Urganci et al., (2022) | Strengths: • The clinical and patient demographic information in HES (for both maternity and further inpatient episodes after birth) allowed for a broader range of maternity care and neonatal outcomes to be compared  • Access to individual data for each woman and infant, rather than national aggregate data, enabled comparison of differences in intervention and outcome rates according to women’s ethnic and socioeconomic backgrounds.  Limitations: • no routine data regarding antenatal care, and therefore it is not possible to directly attribute changes in outcomes to changes that have occurred as a result of the pandemic. |
| Dundas et al., (2023) | Limitations: • we were able to undertake complex data linkages between GUS cohort 2 and routine NHS data in Scotland.   Strengths: • GUS data linked to routinely collected NHS data provide an opportunity to assess longer-term outcomes for children and mothers in a cost-effective and non-burdensome way on participants. |
| Smith et al., (2023) | Strengths: • We were able to have population coverage by combining data from two national datasets |
| Robinson-Smith et al., (2019) | None |
| Cavallaro et al., (2024) | Limitations: • Could only control for characteristics and include outcomes captures in routine data • Diffential linkage of routine data represents inequality • Length of data approval and access  Suggests the following for improving similar evaluations using routine data: • more complete recording of characteristics used to prioritise mothers for enrolement.  • detailed recoding of programme metadata. • Individual-level or aggregate data on characteristics of all mothers-to-be offered enrolment and those who declined versus those who accepted enrolment. • Improved, high-quality data on usual care. |
| Fair and Soltani (2024) | Limitations: • education was poorly documented within the maternity notes. • childhood anthropometric data was collected within routine care and therefore recorded by various personnel, which may limit standardisation. |
| Tassie et al., (2024) | Limitations: • When using MHSDS, several challenges with retrieving the length of hospital admission and cluster assignment (process of classifying patients into care clusters based on their level of need and complexity) data were encountered, such as data ambiguities and missingness. Several data-cleaning assumptions were required.  • Variable availability and data quality. Within this dataset, we were missing four-months of ‘look-back’ data (during the change-over to MHSDS) for previous mental health contacts.  • Other issues related to data availability and quality including a shorter ‘look-back’ period for younger women as they did not have access to child/adolescent mental health care contacts therefore we only had full 10-year look-back data for women aged 28 or over.  Strengths: • Using linked national datasets provided a large number of patient records of care provided by the NHS, providing greater precision in the cost estimates. |
